# “I don’t feel safe sitting in my own yard”: Chicago resident experiences with urban rats during a COVID-19 stay-at-home order

**DOI:** 10.1101/2020.11.25.20238741

**Authors:** Maureen H. Murray, Kaylee A. Byers, Jacqueline Buckley, Seth B. Magle, Dorothy Maffei, Preeya Waite, Danielle German

**Author notes:** Corresponding author: Name: Maureen H. Murray, Address: 2001 N Clark St., Chicago IL, 60614.

## Abstract

**Background:** Encounters with rats in urban areas increase risk of human exposure to rat-associated zoonotic pathogens and act as a stressor associated with psychological distress. The frequency and nature of human-rat encounters may be altered by social distancing policies to mitigate the COVID-19 pandemic. For example, restaurant closures may reduce food availability for rats and promote rat activity in nearby residential areas, thus increasing public health risks during a period of public health crisis. In this study, we aimed to identify factors associated with increased perceived exposure to rats during a stay-at-home order, describe residents’ encounters with rats relevant to their health and well-being, and identify factors associated with increased use of rodent control.

**Methods:** Urban residents in Chicago, a large city with growing concerns about rats and health disparities, completed an online questionnaire including fixed response and open-ended questions during the spring 2020 stay-at-home order. Analyses included ordinal multivariate regression, spatial analysis, and thematic analysis for open-ended responses.

**Results:** Overall, 21% of respondents (n=835) reported an increase in rat sightings around their homes during the stay-at-home order and increased rat sightings was positively associated with proximity to restaurants, low-rise apartment buildings, and rat feces in the home (p≤0.01). Many respondents described feeling unsafe using their patio or yard, and afraid of rats entering their home or spreading disease. Greater engagement with rodent control was associated with property ownership, information about rat control, and lower incomes (p≤0.01).

**Conclusions:** More frequent rat encounters may be an unanticipated public health concern during periods of social distancing, especially in restaurant-dense areas or in low-rise apartment buildings. Rat presence may also limit residents’ ability to enjoy nearby outdoor spaces, which otherwise might buffer stress experienced during a stay-at-home order. Proactive rat control may be needed to mitigate rat-associated health risks during future stay-at-home orders.

## Background

Urban rats create public health risks in cities around the world. Their adaptability to a range of environments has allowed them to thrive in close association with people. This tendency toward close contact with people is concerning for public health because rats can carry many zoonotic pathogens associated with illness in people such as *Leptospira interrogans, Escherichia coli*, and *Clostridium difficile* [1]. Rats can also act as a mental health stressor [2] and cause billions of dollars in property damage [3]. Identifying the drivers of rat encounters in residential areas may help mitigate health concerns. For example, rats typically aggregate in city blocks where food sources such as garbage are abundant [4, 5]. Changes in the availability of these resources for rats in residential areas may therefore alter local rat abundance and rat-associated health risks.

Local environmental changes can impact rat abundance and distributions, and these changes can be influenced, in turn, by global phenomena. In spring 2020, millions of people around the world were required to stay at home to mitigate the COVID-19 pandemic, resulting in altered activity in and around businesses, food establishments, public spaces, and the home. Urban residents in multiple countries reported increased rat sightings during this period, prompting media articles and public health recommendations from the Center for Disease Control [6, 7]. These increases in rat sightings were hypothesized to be the result of restaurant closures and subsequent changes in the distribution of garbage as rats search for new food sources [7]. Indeed, an early analysis found that rat complaints increased in New York City during social distancing restrictions near closed food establishments [8]. Although these results are preliminary, this pattern underscores the potential for surveys to document changes in resident experiences with rats that are not captured in complaint data. For example, residents living near restaurant-dense areas may be more likely to experience increased contact with rats or their excreta. In addition to changes in rat activity, residents may also observe rats because they are spending more time in and around their residence. Regardless of cause, understanding whether residents experienced more frequent or severe encounters with rats during the COVID-19 pandemic will help mitigate additional and unanticipated public health risks associated with rats.

The impacts of rats on residents will also be determined by the context of these interactions. Resident experiences with rats while staying at home may vary depending on local environmental and social factors. Previous studies have demonstrated that rat abundance and rat-associated health risks vary significantly between neighborhoods [5, 9] and is typically higher in low-income areas [10, 11], likely because of building conditions and resources available to control rats. Residents’ encounters with rats might also differ based on their housing conditions; property owners may face monetary burdens from controlling rats while some renters may have negligent landlords. Some residents may also be disproportionately impacted by rat infestations, for example if they have young children, pets, or rarely leave the house. Identifying which communities are most vulnerable to rat infestations will help prioritize proactive rodent control policies and education campaigns to mitigate health risks from rats.

To understand how social distancing policies changed resident encounters with rats, we evaluated how a stay-at-home order influenced three aspects of resident vulnerability to rats: exposure to rats, impacts of rats on human health and well-being, and adaptive capacity to mitigate these interactions [12]. We assessed rat exposure as the frequency of rat encounters and changes to these encounters as compared to before the stay-at-home order; impacts of rats as the frequency and change in rat encounters relevant to human health or well-being; and adaptive capacity by identifying factors associated with engagement with rodent control. We did so by surveying residents in Chicago, a large city with increasing problems with rats [5] and disparities in public health outcomes between neighborhoods [13]. Based on anecdotal reports, we predicted that residents living in proximity to more restaurants and especially in lower-income areas would be more likely to experience an increase in rat encounters. We also predicted that engagement with rodent control would be positively associated with increased rat sightings, motivation (i.e. concern about rats), and capacity (i.e. information and income). Understanding resident experiences with rats during the stay-at-home order will help further understand the impacts of rat infestations on human health and well-being as well as anticipate community health issues as cities re-open or restrict activities to control the current pandemic.

## Methods

A cross-sectional study design was used to collect information from Chicago residents about their experiences with rats during the stay-at-home order. The survey was formatted to collect responses through SurveyMonkey, an online survey service to ensure rapid distribution of surveys, which would have been impossible using paper surveys. The survey was available online between April 27 and June 6, 2020 corresponding with Chicago’s stay-at-home order, which was in effect from March 21 to June 3, 2020. During the stay-at-home order, all residents were requested to stay at home except for essential needs and restaurants could not provide dine-in service [14].

To obtain responses from all Chicago neighborhoods, we distributed the link to our online survey via email to all 50 Aldermanic offices (i.e. elected officials who represent city wards) and at least one community organization in all 77 community areas. Prior to distributing the survey, a list of potential questions was pilot tested (n = 21) and refined to improve comprehension. The survey was available and advertised in English and Spanish. Survey respondents were deemed eligible if they were over the age of 18 and had lived in their current residence for at least six months. To increase the accuracy of responses, survey questions were preceded by an introduction that distinguished the appearance and sign (e.g. droppings, burrows) of rats relative to mice. All survey participants anonymously provided written informed consent via checkboxes prior to taking the survey. The Lincoln Park Zoo Institutional Review Board approved our informed consent protocol and deemed this study exempt from the requirements of 45 CFR 46.

The survey questionnaire (Supplementary file 1) was designed to follow the methods of German and Latkin [2]. The survey questions aimed to describe any changes in residents’ exposure to rats (i.e., frequency and context of rat encounters), the impacts of rats on their health or well-being (i.e., zoonotic exposure risk and mental health impacts); and their adaptive capacity to addressing rats (i.e., changes in behaviors including use of rodent control). Within each topic, we asked respondents to report the *frequency* of an event in the past month from “never” to “daily or almost daily” and we asked about any *change* relative to a month prior to taking the survey. Respondents also had the opportunity to describe their feelings and experiences associated with rats in their own words (see below).

We hypothesized that resident experiences with rats would be mediated by several demographic and environmental factors. To understand these relationships, we asked respondents to self-report their age group, gender, children in the household (yes/no), type of housing, whether they were renters or property owners, time spent outside the house per week, their neighborhood, and their closest major intersection. For analysis, we grouped housing types into single-family homes, low-rise multi-unit buildings (<10 units), and high-rise multi-unit buildings (≥ 10 units; [15, 16]. We also included the median household income of the respondent’s census tract [15] if they provided their closest major intersection.

### Exposure

To determine whether residents experienced more frequent rat encounters during the stay at home order (i.e. exposure), we asked respondents to report the frequency and change in rat sightings at two geographic levels: in/around their home; and on their city block. Because rat abundance is known to increase during the spring [5, 8], and because our survey coincided with this time period, we also asked about their change in rat sightings in spring 2020 relative to previous years.

To test the hypothesis that rat activity would increase in areas near restaurants during the stay-at-home order, we undertook a spatial analysis. To do this, we compared respondents’ reported change in rat sightings with the number of restaurants within 500m of their closest major intersection. We chose a buffer size of 500m because rats have small home ranges on the order of tens to hundreds of square meters [17] and the closest major intersection may be several blocks away from the respondent’s residence. We accessed the location of all food vendors in Chicago using food inspections data from the Chicago Department of Public Health’s Food Protection Program [18]. We treated the count of restaurants within 500m as a continuous variable. However, the count of restaurants was right skewed (median = 51, mean = 64, range = 1 - 425) and so we removed outliers and log-transformed the values prior to analysis.

We also accessed municipal rat complaint data to complement survey responses about any increase in rat infestations. The city of Chicago records public complaints about rats made via 311 calls and online. In response to complaints, city managers will distribute rodenticide bait in the complainant’s alley. Rat complaint data can be a useful way to measure the timing or locations of rat infestations [5, 8, 19]. Although rat complaints are not a direct measure of rat populations, the biases associated with complaints (e.g. knowledge of the 311 system) may differ from sampling bias in survey research (e.g. internet access) and therefore similar trends in these complementary data sources may support survey results. We accessed rat complaints made to the city between January 1 and June 6, 2020 through a Freedom of Information Act (FOIA) request to the City of Chicago Department of Streets and Sanitation. We then calculated the change in 311 rat complaints made in each census tract in March 2020 relative to May 2020 as these were the time periods relevant to our survey. All spatial analyses were performed in QGIS 2.18.14 [20].

### Impacts of rats

To understand the impacts of rats on public health risks, we asked respondents to report the frequency and change in rat encounters that may lead to the transmission of zoonotic pathogens such as touching rat feces or being bitten by a rat. We also asked respondents to describe how they felt about their interactions, including their concerns about these encounters.

### Adaptive Capacity

To assess adaptive capacity, we asked residents about changes in their behaviors during the stay-at-home orders that were specifically meant to minimize contact with rats. We also included questions about resident engagement with rodent control as a measure of adaptive capacity. To test whether an increase in rat sightings during the stay-at-home order was associated with increased engagement with rodent control, respondents were asked about the frequency and change in their use of rodent control such as reporting rat complaints to the city and/or calling a pest professional. We hypothesized that engagement with rodent control is mediated by motivation and ability. We thus asked if respondents were more concerned about rats now than they were a month ago and if they had enough information to control rats on a 5-point Likert scale from “strongly disagree” to “strongly agree”.

### Thematic analysis

We analyzed respondents’ open-ended responses to the question “If you currently have rat problems or had them in the past, how did that make you feel?” using a “thematic framework” [21, 22]. A preliminary coding framework was developed by MM, JB, PW, and DM for thematic analysis of the responses. Some codes (i.e. emotions associated with rats) were determined a priori based on the survey question. Many other codes emerged organically based on respondents’ tendency to discuss certain topics (e.g. how rats impact their day to day life). The survey responses were coded manually using the program Dedoose [23] and all responses were coded by at least two reviewers. The thematic framework was revised once all responses were coded. Themes were developed based on the objective of the question (i.e. describe respondents’ feelings about rats) and by identifying concepts that were mentioned by multiple respondents in relation to the study objectives.

### Statistical analysis

For quantitative analysis of survey questions, we used generalized linear models to identify variables significantly associated with exposure to rats (i.e. change in rat sightings), impacts of rats (i.e. change in contact with rats or rat feces), and adaptive capacity (i.e. engagement with rodent control). For all of these three outcomes, the response variables were rating scales with the categories “more often”, “about the same”, and “less often” relative to a month prior. Because the response variables of interest were ordered categories, we used ordinal regression in R using the package MASS [24, 25] and the reference category for all three outcomes was “more often”. We included the demographic and socioeconomic variables listed above such as housing type and income in all models. We added additional predictor variables to the models based on the factors we hypothesized would be associated with a change in rat sightings (i.e. number of restaurants within 500m), change in contact with rats or rat feces (i.e. change in rat sightings), and engagement with rodent control (i.e. change in rat sightings, concern about rats, information about rats). All continuous variables were centered and scaled prior to analysis.

## Results

### Survey Participant Characteristics

The online survey was shared electronically by nine community organizations and by the aldermanic offices in 29 of Chicago’s 50 wards. In total, we received 835 at least partially completed surveys from eligible respondents but none in Spanish. Of the 672 respondents who provided demographic data, 67% (N = 453) identified as female, 31% as male (N = 205), 1% as non-binary (N = 5), and 1% preferred not to say (N = 9). Because a small proportion of respondents self-identified as non-binary, we were unable to include their responses when including gender as a covariate in generalized linear models. All age groups were represented although there were fewer respondents in the 18-24 category (3.5%) relative to other categories (13% - 19%, Table S1). Relative to Chicago’s population, respondents were disproportionately more likely to self-identify as female (51% vs 67%, respectively) and property owners (45% vs 66%), but there was no significant bias in age class (Table S1, see Supplementary file 2). Most respondents indicated that they had heard of the survey from their Alderman/city council member (47%), from a community organization (17%) or from social media (17%).

Of the eligible respondents, 740 provided their neighborhood and 627 provided their closest major intersection. We received responses from 106 neighborhoods, mostly in the North Side (Figure 1). We received more responses from community areas with higher incomes and with more 311 rat complaints during the stay-at-home period (β_Income_ = 8.36 ∓ 2.39 SE, t = 3.49, p = 0.001; β_Complaints_ = 8.61 ∓ 2.31 SE, t = 3.73, p = 4.82 x 10-4; model adjusted R2 = 0.43, F(2,50) = 20.63, p = 2.93 x 10-7). Based on census tract data, the average median household income for respondents who provided their closest major intersection was $82,888 ∓ 34,182 (range: $16,953 - $168,352).

**Figure 1:**
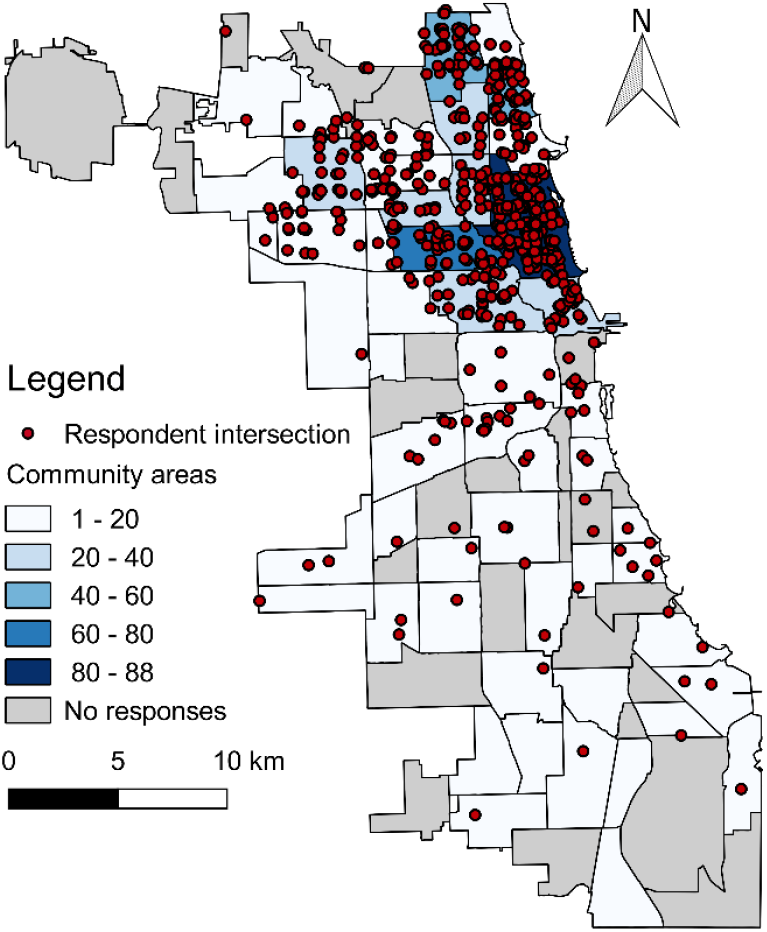
Map of Chicago showing community area boundaries and the locations of survey respondents. Community areas are shaded based on the number of respondents who self-reported their neighborhood of residence (n = 740). Red circles indicate the closest major intersection reported by respondents (n = 627). The locations of respondent intersections were offset by a random distance within a 500m buffer to maintain respondent privacy.

We received 485 responses to the open-ended question “If you currently have rat problems or had them in the past, how does that make you feel?” These respondents were more likely to report seeing rats at least once in the past month but otherwise did not differ significantly from the other respondents (Data not shown). Based on a thematic analysis of these responses, four themes emerged: Context of rat encounters, Emotional responses to Rats; Impacts of rats; and Accountability and responsibility for rodent control (Figure 2). We will discuss these themes as they relate to our study objectives below.

**Figure 2:**
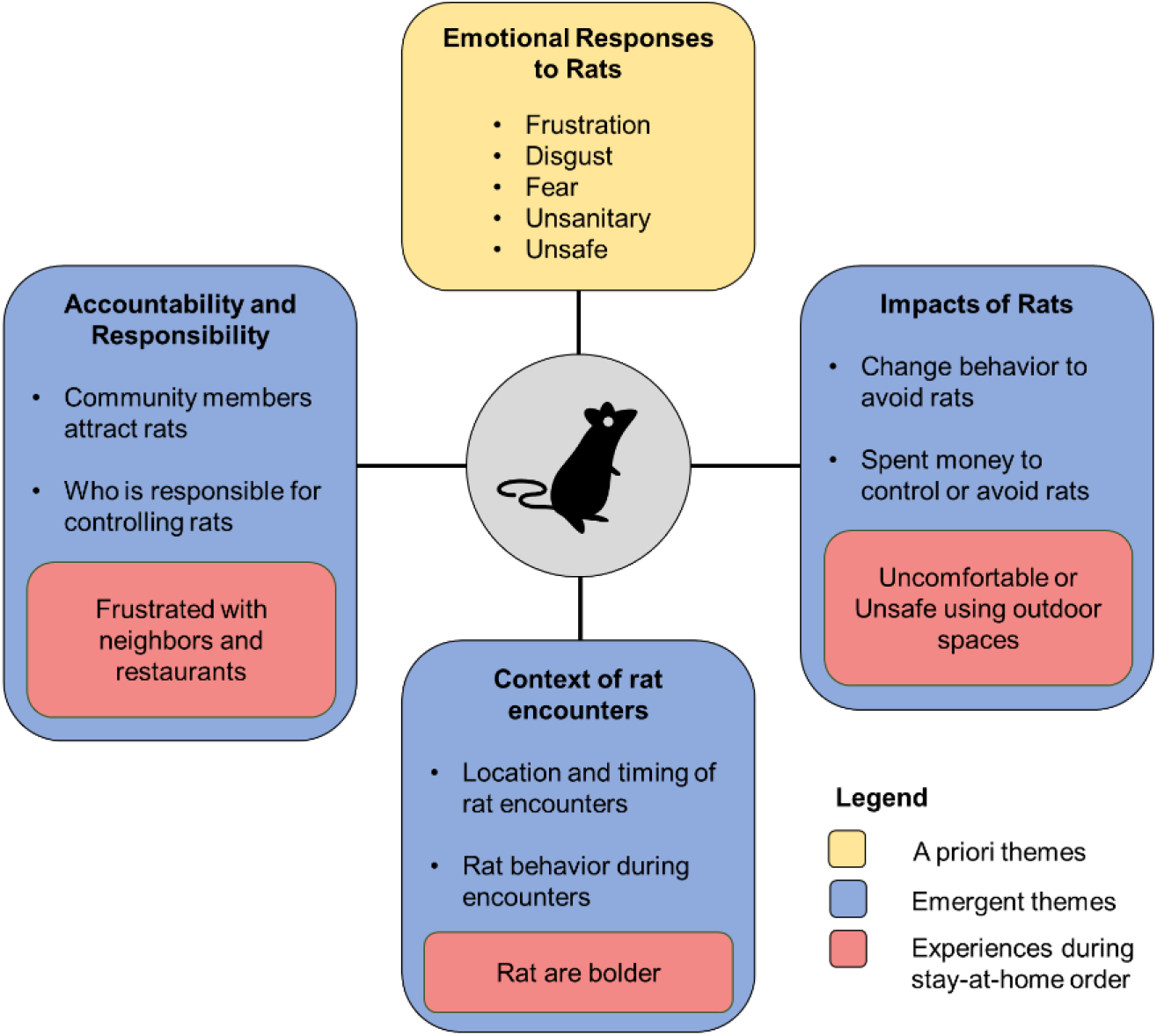
Thematic structure describing resident experiences with rats during the stay-at-home order. Yellow and blue squares represent major themes identified in open-ended survey responses using thematic analysis. Red squares represent experiences unique to the stay-at-home order period.

### Exposure to rats: “there is a bigger issue now”

Of eligible respondents, 21% observed more rats in or around their home and 23% observed more rats on their block during the stay-at-home order (Table 1). Conversely, half as many respondents observed fewer rats around their home (11%) or on their block (10%) during the stay-at-home order (Table 1). Respondents were more likely to observe an increase in rats on their block if they lived near more restaurants (p = 0.001), spent more time outside (p = 0.001), and lived in a small multi-unit building (p = 0.003) (Table 2, Figure 3). Respondents who observed an increase in rats were also more likely to report more rats in 2020 relative to previous years (p < 0.001; Table 2). Respondents were also more likely to observe greater numbers of rats in or around their homes if they lived in a small multi-unit building (Table S2). We found no significant correlation between an increase in rat sightings from survey responses and increased 311 complaints in the same census tract (Table 2 and Table S2).

**Table 1:** Frequency of rat sightings and encounters during March - June 2020 stay-at-home order in Chicago, Illinois, USA. Residents were asked to recall the frequency of experiences during the past month and compare their current experiences with a month ago.

|  | Frequency during quarantine |  |  |  | Change during quarantine |  |  |  |
| --- | --- | --- | --- | --- | --- | --- | --- | --- |
|  | Never | Rarely (Less than weekly) | Frequently (Weekly) | Daily or almost daily | Does not occur | Less often | About the same | More often |
| Rats in home | 253 (30%) | 260 (31%) | 219 (26%) | 109 (13%) | 173 (21%) | 8 (11%) | 400 (48%) | 173 (21%) |
| Rats on block | 186 (22%) | 243 (29%) | 263 (31%) | 145 (17%) | 130 (16%) | 84 (10%) | 423 (51%) | 195 (23%) |
| Saw chewed objects | 498 (69%) | 139 (19%) | 58 (8%) | 24 (3%) | 266 (37%) | 95 (13%) | 271 (38%) | 90 (13%) |
| Rat feces in home | 453 (63%) | 154 (21%) | 80 (11%) | 31 (4%) | 264 (37%) | 98 (14%) | 261 (36%) | 98 (14%) |
| Touched rat feces | 661 (91%) | 52 (7%) | 16 (2%) | 1 (0.1%) | 480 (67%) | 83 (12%) | 144 (20%) | 13 (2%) |
| Touched a rat<br>(alive or dead) | 650 (89%) | 70 (10%) | 9 (1%) | 0 | 504 (70%) | 91 (13%) | 118 (17%) | 7 (1%) |
| Bitten by rat | 725 (99%) | 3 (0.4%) | 1 (0.1%) | 0 | 529 (74%) | 80 (11%) | 108 (15%) | 2 (0.3%) |

**Table 2:**
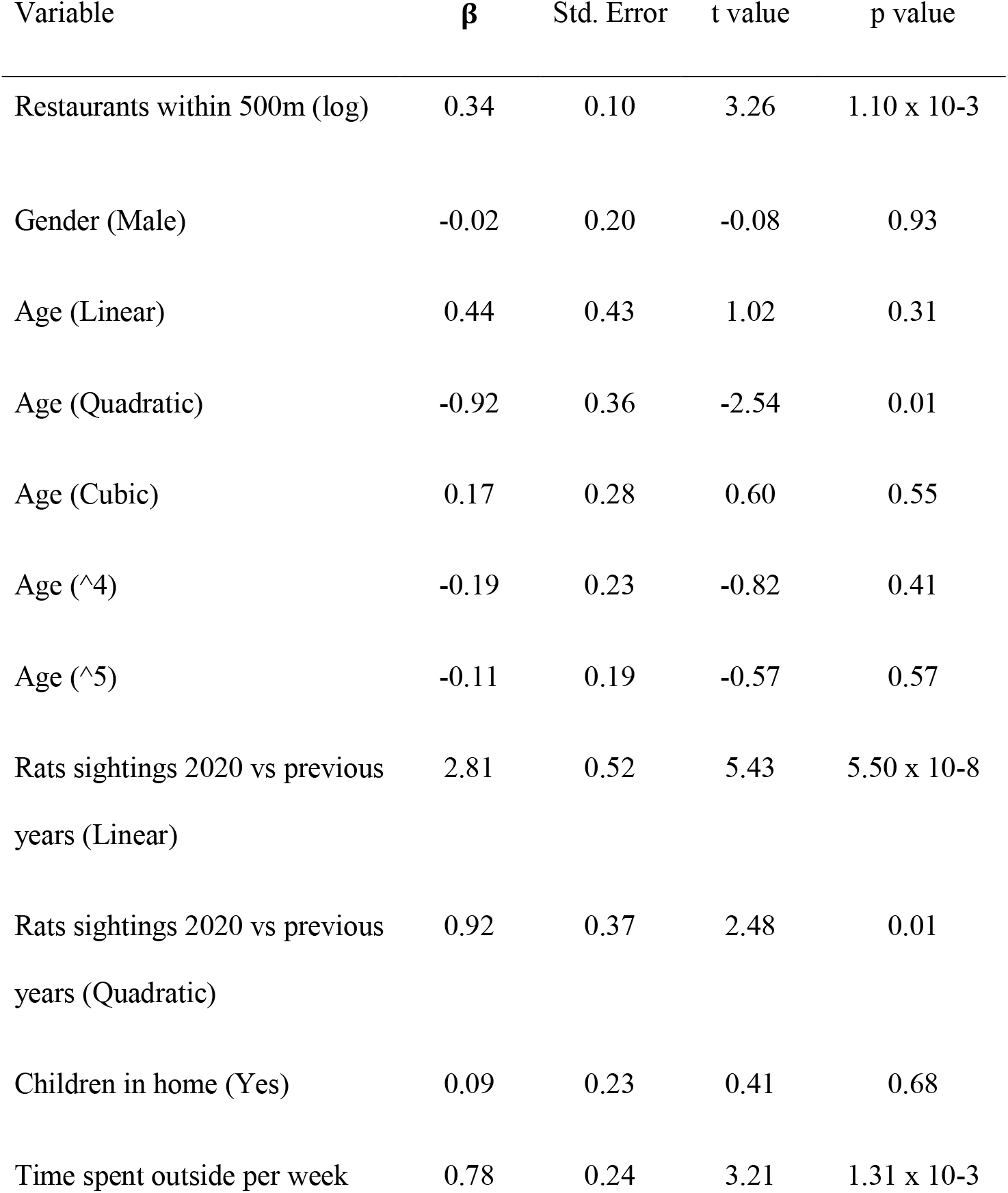
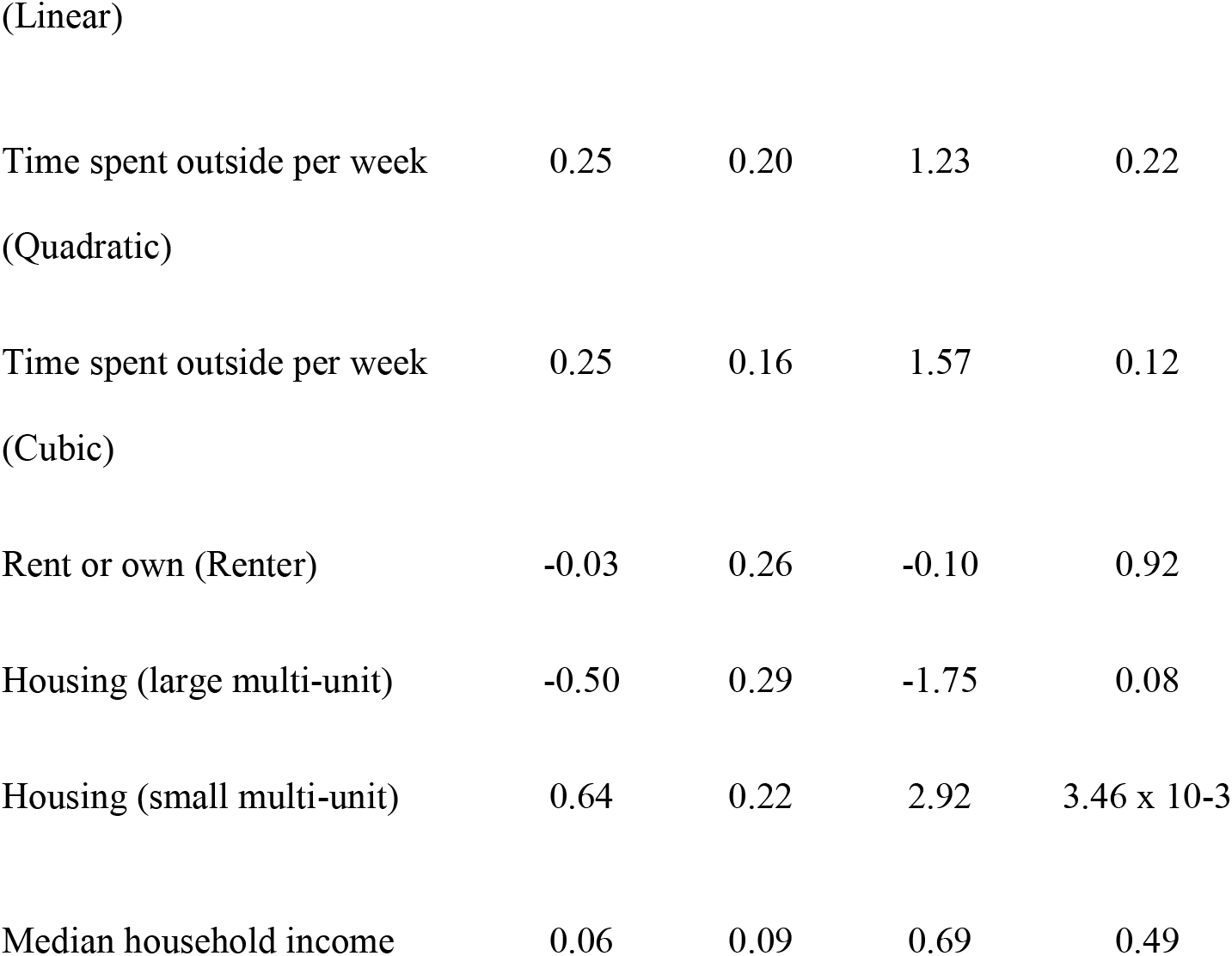
Ordinal regression output for variables hypothesized to be associated with a change in rat sightings on respondents’ block of residence during quarantine. The response categories were “more rats”, “about the same”, “fewer rats”, and “I see no rats here” and the reference category was “more rats”.

**Figure 3:**
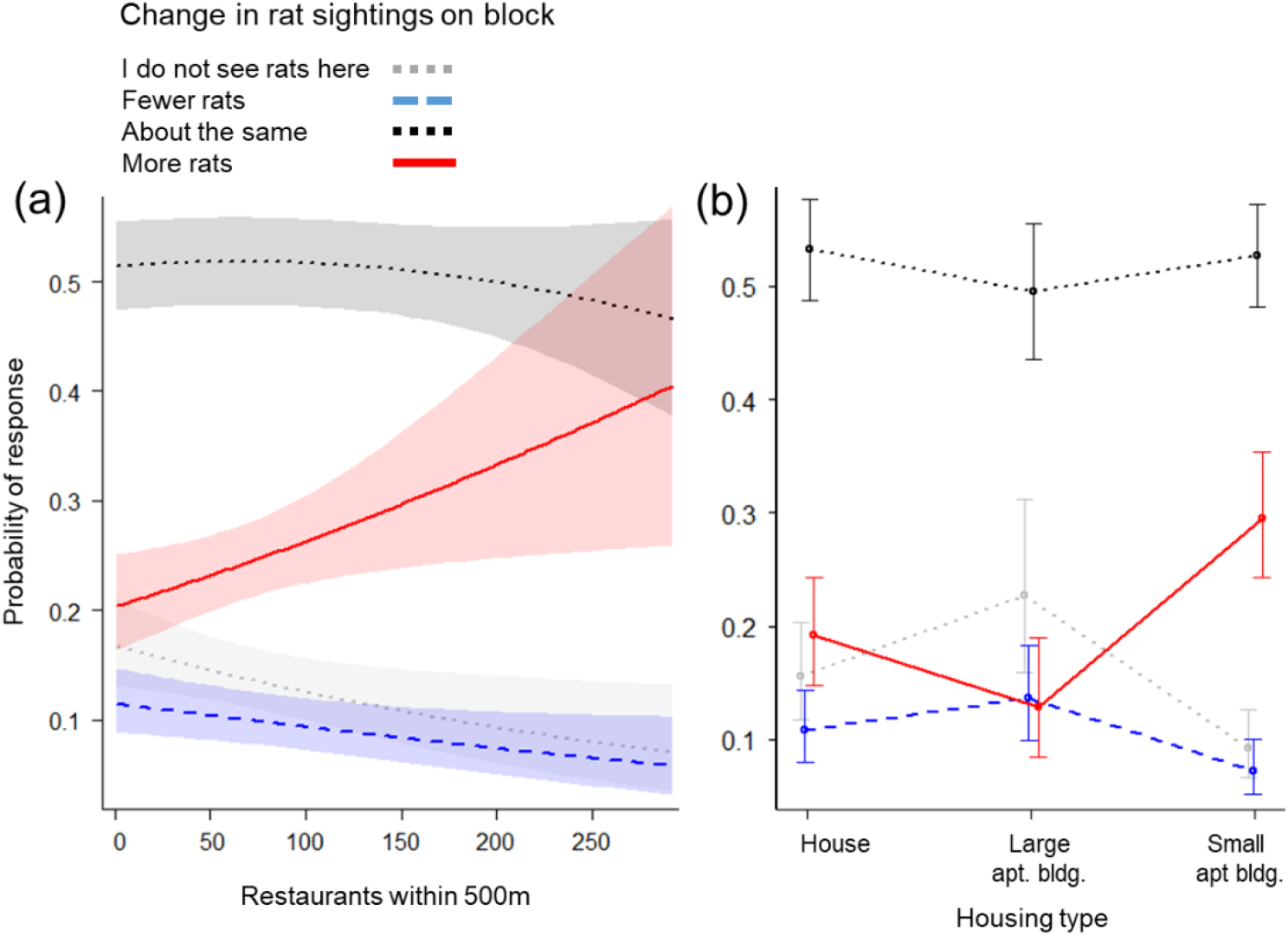
Relationships between the change in rat sightings during the stay-at-home order in Chicago and proximity to restaurants (a) or housing type (b) based on ordinal regression of survey responses. Lines show the probability of a survey respondent reporting more rats (red solid line), fewer rats (blue dashed line), about the same number of rats (black dotted line) or no rats (gray dotted line) relative to a month prior. The count of restaurants was log-transformed for analysis due to right skewness and back-transformed for ease of interpretation. Shaded bands (a) or error bars (b) show 95% confidence intervals.

In their open-ended responses, several residents mentioned that they see more rats now that restaurants have closed. Most surmised that there was less garbage available to rats following restaurant closures, which caused rats to move into residential areas.

*“as a landlord, It’s disgusting to have rats on our property despite our efforts to keep everything clean. The restaurants we share an alley with are mediocre at best in keeping trash off the ground. Now with less to no food in the dumpsters, the rats are coming & burrowing in our yard”*

Several respondents also mentioned that rats appeared to be less afraid of people during the stay-at-home order. These respondents described rats being more visible and active during the day, potentially because rats were searching for new food sources after restaurant closures.

*“Rats are a normal part of city life and usually keep to themselves and the alleyways, but since restaurant closings I’ve noticed them getting “bolder”, often finding them crossing yards and porches in daylight and walking very close. I suppose because with the lack of restaurant garbage their food sources have disappeared?”*

### Impacts of rats: “Unsafe in my own home”

Several respondents reported interactions with rats of public health concern. In the month prior to taking the survey, 11% of respondents had touched a rat, 10% touched rat feces, and four respondents (0.5%) were bitten by a rat at least once (Table 1). Although over 70% of our respondents resided in the North Side of Chicago, three respondents that reported being bitten by a rat lived in less-affluent South Side neighborhoods, representing 5.0% of all South Side respondents. Respondents who observed an increase in rats in their home were also more likely to observe increased rat feces in their home (p < 0.01; Table 3; Figure S1).

**Table 3:**
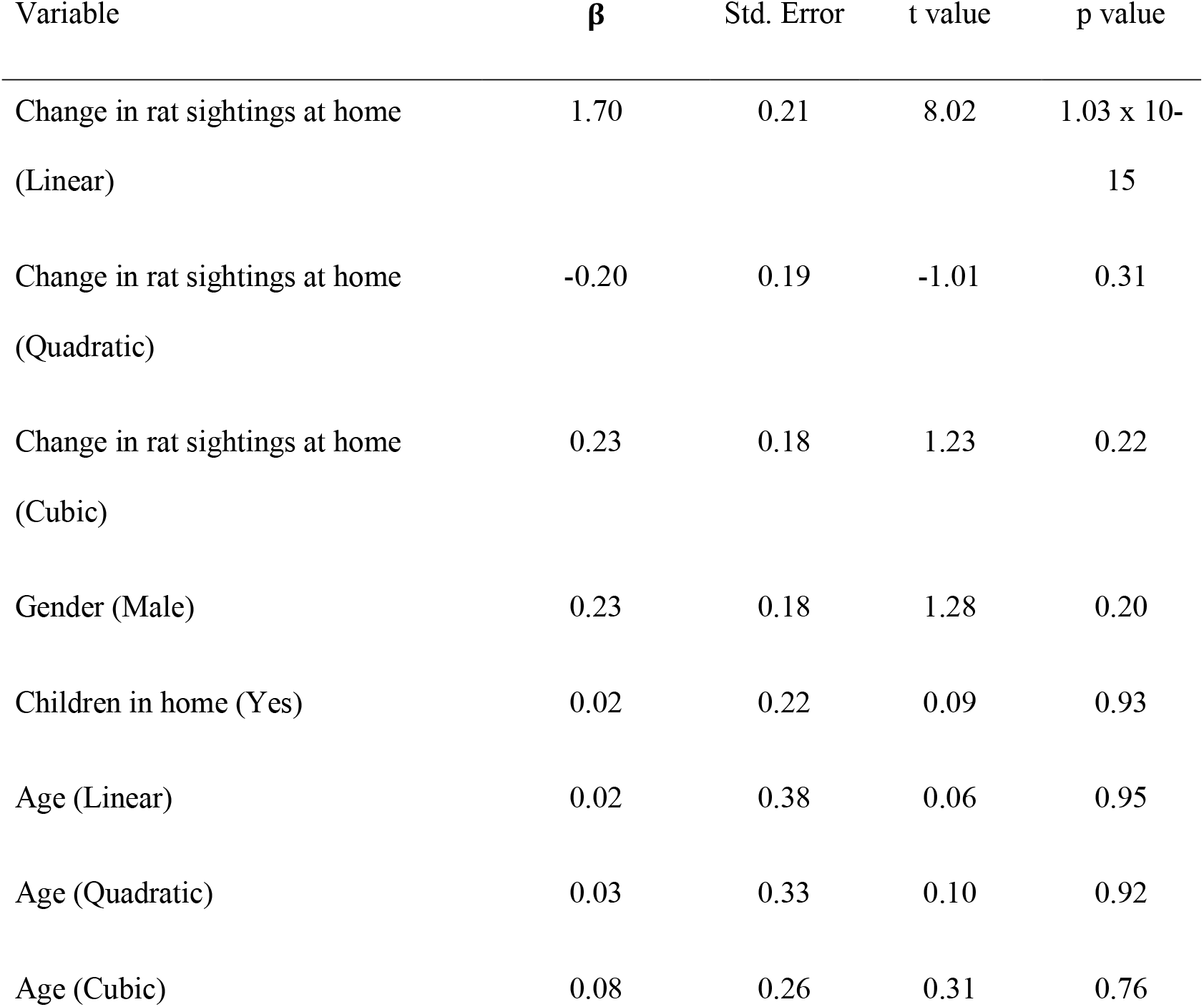
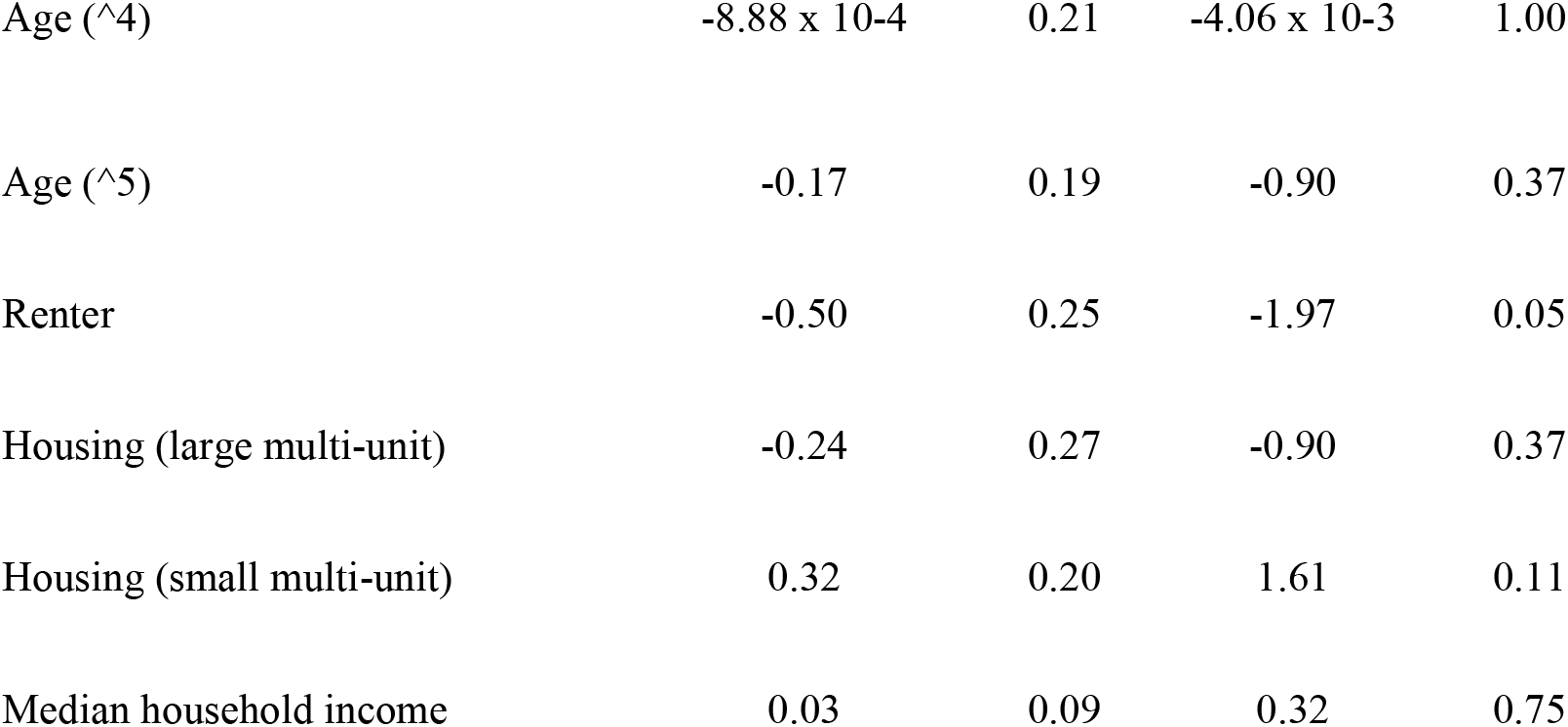
Ordinal regression output for variables hypothesized to be associated with a change in rat feces in respondents’ homes during quarantine.

Most respondents described feeling concerned/worried (21% of respondents), afraid (18%), unsanitary (17%), disgusted (14%) or frustrated (14%) in the context of rat encounters. Many respondents described feeling unsanitary, dirty, or unclean because of rats living in their home. Of respondents who expressed feeling frustrated, most were frustrated with their neighbors or other community members for actions they believe attracted rats or were frustrated at the difficulty of eliminating rats (see below).

*“A home violation. It feels disgusting. I’m concerned for my kids and my home. I’m scared they’ll come on to us when we sleep*.*”*

Many respondents mentioned being concerned for their family’s health or safety because of the risk of disease transmission or aggression from rats. Specifically, many respondents mentioned being concerned about their children playing in areas with rats or rat feces or about their pets being exposed to diseases from rats or rat poison.

*“I have a nice home, and seeing rats in and around it brings me so much anxiety, it’s hard to put into words. It makes me feel dirty and scared. I am afraid my cats, my kids, or I will be bitten and hurt, or that they will bring disease to my home*.*”*

Residents who reported observing more rats during the stay-at-home order also described feeling frustrated about rats or worried about rats entering their home more frequently than other respondents. These respondents were also the only group who mentioned feeling unsafe because of rats or being concerned or afraid about the risk of disease from rats.

*“It makes me feel unsafe from disease*.*” “I feel scared that my family can get sick*.*”*

Another emergent theme in the open-ended responses was changes in resident behavior to avoid encountering rats. Many respondents mentioned feeling uncomfortable using their outdoor spaces such as patios or yards because they were afraid of encountering rats or disgusted by the presence of rat droppings. Several respondents emphasized their discomfort with recreating outdoors after dark when rats are most commonly active.

*“I fear going out at night as I often encounter rats running through the yard and in the street. I am unable to use my back patio after dark*.*”*

When describing this discomfort, respondents often mentioned feeling unsafe or unwelcome on their own property. These feelings were often shared in association with perceptions that parties responsible for pest control were being neglectful.

*“Rats make me feel like I’m not welcome in my own backyard even though it’s my yard. It’s very frustrating when building neglect and slum lords ignore their properties and rats take over under garages and buildings. It makes me feel unsafe for my family as we see rats daily in our backyard*.*”*

*“every evening a rat runs across our backyard and sometimes in our gangway. I don’t feel safe sitting in my own yard. we have children and it feels unsafe and unclean for them even though we maintain cleanliness in/out of our house. they are scary and i’m afraid they may become aggressive if hungry*.*”*

Several residents highlighted that this discomfort with using private outdoor spaces because of rats felt especially limiting or frustrating during the stay-at-home order because they had few options to safely leave their home or recreate outdoors.

*“[I feel] Limited as I don’t enjoy being in my yard. My quality of life is greatly affected”*

### Adaptive capacity to control rats: “I can’t do this by myself”

Respondents were more likely to report using more rodent control during the stay-at-home order if they observed increased rat sightings in their home (p < 0.001), if they were more concerned about rats than they were previously (p < 0.001), if they were property owners (p < 0.001), if they had enough information about rats (p = 0.03), and if they had lower incomes (p = 0.01; Table 4). Of respondents who reported observing rats daily, 42% never called 311 and of respondents who reported observing rats weekly, 59% never called 311. Respondents were significantly more likely to call 311 at least once in the past month if they observed an increase in rats in their home (β = 1.77 ± 0.44, p < 0.01), if they were more concerned about rats than they were previously (β = 1.85 ± 0.40, p < 0.01), if they strongly agreed that they had enough information to control rats (β = 1.27 ± 0.37, p < 0.01) and if they were property owners rather than renters (β = −1.14 ± 0.40, p < 0.01; Table S3). These same variables were significantly associated with calling a pest professional at least once in the past month (Table S4).

**Table 4:**
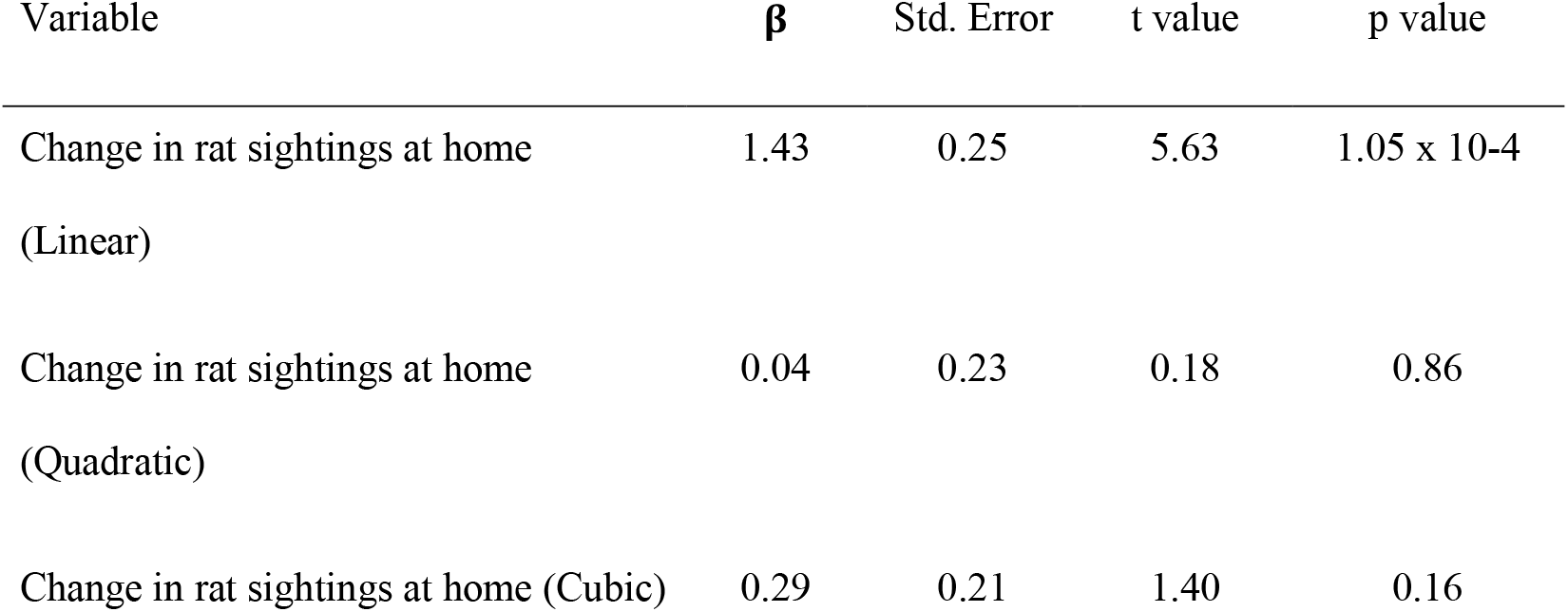
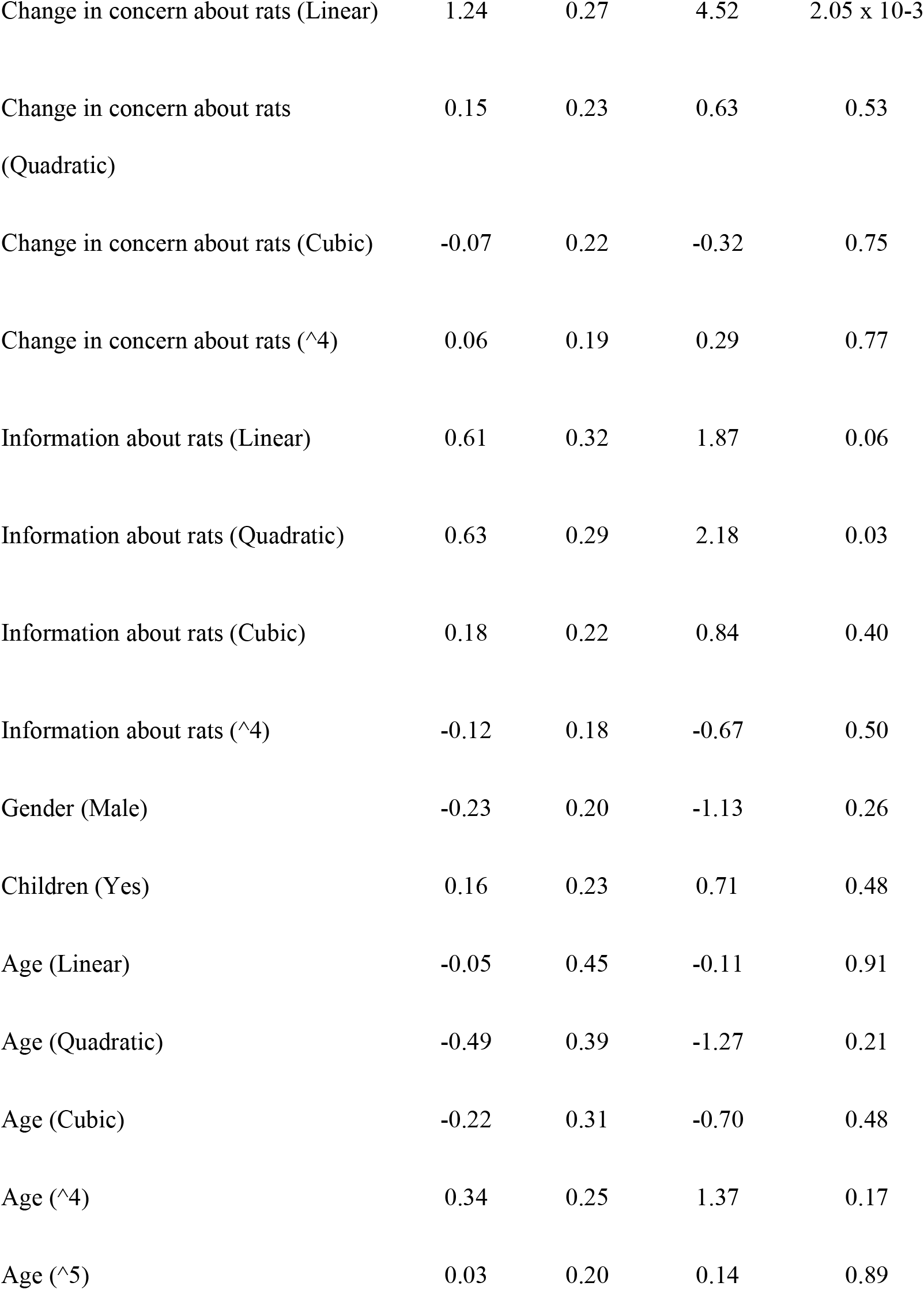
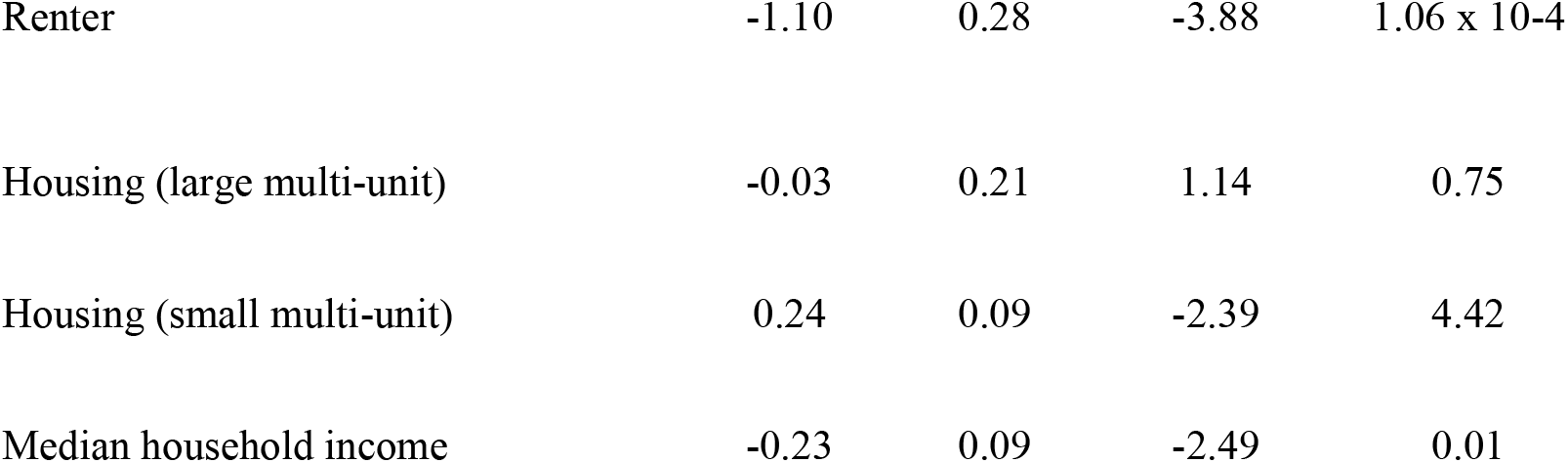
Ordinal regression output for variables hypothesized to be associated with a change in engagement with rodent control during the quarantine period.

A common theme that emerged from respondent descriptions was concerns about who is responsible for rat problems and accountable for rodent control. Many respondents expressed that their neighbors attracted rats, often by improperly managing garbage in alleys, or were not active enough in controlling their rat problems. Many respondents also expressed that the city government was ultimately responsible for rodent control and could be more active in their neighborhood.

*“Note that since the pandemic the neighborhood is getting trashed. People are not cleaning after their dogs like they used to n are throwing garbage on sidewalks n lawns. Is this part of their rebellion against being told what to do ie stay inside, wear masks, don’t congregate? Because in this area all of that is blatantly ignored. So I feel frustrated that these new habits encourage rats*.*”*

Several others pointed out that they feel alone in dealing with rats and that rats require a community approach for effective rodent control. Many respondents emphasized that they take multiple measures to control rats but they still see rats on their property because of rat burrows or food sources on their neighbor’s property.

*“I worry about the health and safety of my family but I can’t solely solve this issue as the rats burrows are not in my yard but they are simply using my yard as a highway between. This is a neighborhood problem that requires everyone including the city to be involved to solve*.*”*

Indeed, many respondents mentioned how difficult it is to eradicate rats from their property. Many described spending money on multiple control methods that did not always succeed in reducing rat populations. In these cases, many expressed hopelessness that rats can be eradicated.

*“The rats in Chicago seem to be smarter than any pest control measures. I’ve tried snap and glue traps, poison, and city services. I’ve cleaned regularly, made repairs and attempted to seal access points. Evidence of rats still returns*.*”*

One resident also mentioned that they were unable to manage rat harborage because they were taking care of others during the COVID-19 pandemic, leading to increased rat problems during the stay-at-home order.

*“Each spring we do a major clean up. I was a bit delayed this year due to COVID/ Stay at home (had to take care of others). My neighbor started seeing multiple rats in my front yard. Exterminator came out. Confirmed 4 nests. We are choosing to try to take care of by ourselves. 1st step: cleared out all vines, ivy, plants. Discovered 4 HUGE dead rats in addition to the nests*.*”*

## Discussion

In this study, we evaluated whether a stay-at-home order increased resident vulnerability to rat infestations. We found that residents living in proximity to restaurant-dense areas or in small apartment buildings were more likely to report an increase in rat sightings. Residents expressed feeling frustrated with neighbors, restaurants, and restaurant closures for attracting rats. We also found several types of public health risks from rats during the stay-at-home order, including increased exposure to rat feces in the home and feeling unsafe using outdoor spaces. Residents who observed more rats during the stay-at-home order did increase their use of rodent control, however this was more likely for residents with adequate information about rats and lower incomes. Our results suggest that some urban communities may be particularly vulnerable to rat infestations during this stressful and challenging time (Figure 4).

**Figure 4:**
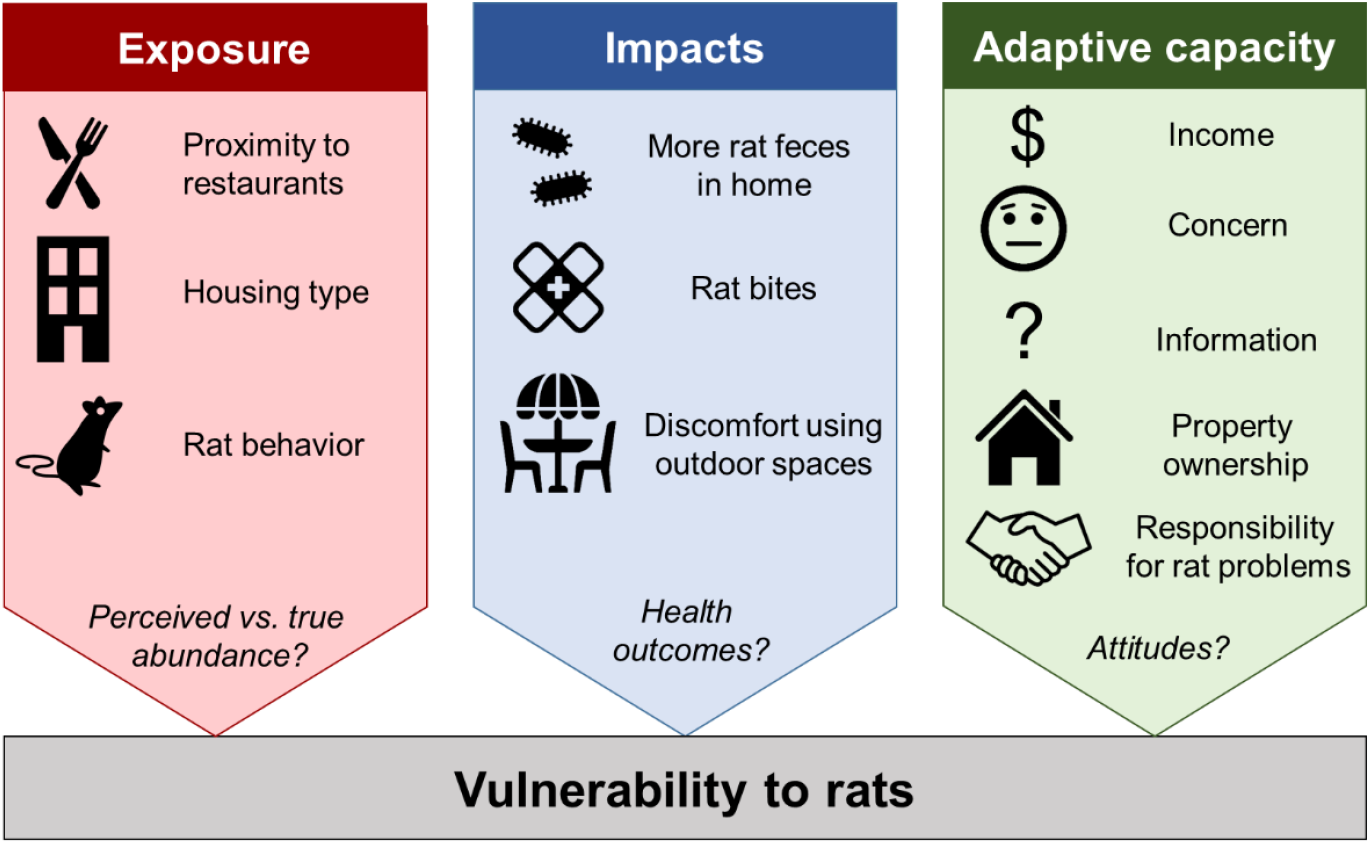
Conceptual framework and summary of main factors found via quantitative and qualitative analysis of survey responses to affect resident exposure to rats (red), impacts of rats on human health or well-being (blue), and adaptive capacity to mitigate rats through engagement with rodent control (green). Text in italics highlights areas for future research.

Although only a subset of respondents observed rats more frequently during the stay-at-home order, increased rat problems appear to be predictable based on their location and type of residence. Respondents living near restaurant-dense areas were more likely to report an increase in rats because of shifts in garbage availability, supporting trends in rat complaints in other cities [8]. With restaurant closures, garbage would have been less plentiful in commercial dumpsters and, simultaneously, food garbage was likely more abundant in residential areas because residents were eating at home more often. Residents in small apartment buildings were also more likely to report increased rat sightings. Although the mechanism driving this pattern is less clear, these residents share walls and garbage facilities with their neighbors who may vary in their fastidiousness with respect to rats. To prevent rat infestations in shared apartment buildings in restaurant-dense neighborhoods, municipalities could prioritize these areas for proactive rodent control including monitoring, rat abatement, and replacing damaged garbage containers [4]. Residents should also be especially vigilant about properly containing garbage and reporting rat sightings to managers. Indeed, many survey respondents emphasized the need for all community members to be actively involved in rat mitigation. Public education campaigns emphasizing rats as a community issue may help promote proactive rat management.

We also found unequal health risks from rats during the stay-at-home order. Respondents who reported increased rat sightings were also more likely to report increased rat feces in the home. Residents in small apartment buildings in restaurant-dense areas may thus be especially at risk for rat-associated zoonotic exposure. Many respondents also reported exposure to rat urine. For example, one respondent stated that “odor from rat urine” made their home office uninhabitable as a result of past rat problems. For many Chicago residents, home offices became primary work locations during the stay-at-home order. Home environments that contain rat feces or urine can expose residents to a variety of zoonotic pathogens such as *Leptospira interrogans*, Seoul hantavirus, and infectious organisms such as *Escherichia coli* and *Salmonella* spp. [9, 26].

Perhaps more concerning, we found that rat bites were mainly reported from Chicago’s South Side neighborhoods. One respondent who reported a rat bite described themselves as “Petrified” and another sought medical attention because of this contact with rats. Although only four respondents reported rat bites, we received disproportionately fewer responses from the South Side, highlighting the need for more information about resident experiences with rats from all Chicago areas. This region of the city is notorious for lower incomes and poorer economic prospects [27], and 93% of South Side residents are Black or African American [28]. These results thus contribute to a growing awareness that multiple aspects of the pandemic have disproportionately impacted Chicago’s Black communities [29].

In addition to public health risks from potential zoonotic exposure, rat infestations during the stay-at-home order may negatively impact resident well-being by restricting their activities. For many Chicago residents, the stay-at-home order caused dramatic changes to their daily routine and use of their home. Staying at home can have negative impacts for resident mental health, such as increased depressive symptoms, symptoms of Generalized Anxiety Disorder, and acute stress [30]. While staying at home, household features such as access to a private yard or patio become more important as a determinant of health and well-being [31]. For example, access to outdoor spaces may help to mitigate the negative impacts of staying at home on residents’ mental health by providing safe sociable spaces [32]. Many respondents described feeling uncomfortable or unsafe using outdoor spaces out of fear that they would encounter rats or come in contact with rat droppings. For those residents who are negatively impacted by staying at home and experiencing rat problems during the stay-at-home order, the relief that private outdoor spaces can offer may be reduced. Controlling rat populations may therefore mitigate physical and mental health risks for residents during this challenging time.

Although residents with increased rat problems were more likely to increase their use of rodent control, knowledge and income appear to be important mediating factors. The city of Chicago receives tens of thousands of rat complaints per year [5]. However, most survey respondents did not report complaints about the rats they saw during the stay-at-home order, potentially because they are not aware of the risks from rats, aware of the 311 rat complaint program, or are not aware that the city will abate rats in their alley for free. The high proportion of respondents who did not report rat complaints might also explain the lack of correlation between our survey results and trends in 311 rat complaints. Beyond the need for information about rats, residents might not engage with municipal rodent control if they do not believe the city will act in response to their rat complaints. Surprisingly, increased use of rodent control was associated with lower incomes, even while controlling for the frequency of rat sightings.

Residents in lower income neighborhoods may be more likely to take on the responsibility and cost of rodent control themselves rather than rely on neighbors or the city. In Baltimore, low-income residents in perceived problem rat areas had less confidence that their neighbors or the city would address rat infestations [2]. Regardless of cause, rat control could impose a monetary burden for some communities during a period of record high unemployment [33] and economic disruption [31]. Public education programs and greater understanding of resident attitudes toward rodent control is therefore needed to increase participation with free municipal services.

Our results suggest avenues for future research to understand the impacts of rat infestations on resident health and well-being. Of note, our sample only included residents with internet access and residents with rat issues may have been more motivated to participate in the study. We received a disproportionately high amount of responses from the North Side of Chicago, potentially because of the aforementioned factors. Future surveys that are mailed to randomly sampled households may provide more robust estimates of rat infestations across the entire city. Importantly, with our survey, it was difficult to determine whether respondents observed a true increase in rat abundance or a perceived increase because residents were spending more time at home. Residents who spent more time outside in their neighborhood were more likely to report increased rat sightings in or around their home (Table S3), suggesting that sightings were likely not only driven by time spent indoors. Future studies that link survey responses with local rat abundance could tease apart actual and perceived risks from rats in urban neighborhoods. Such studies could also document health outcomes associated with rats, which we were unable to examine due to other health concerns during the pandemic.

## Conclusions

Our results suggest that more frequent rat encounters may be an unanticipated public health concern during periods of social distancing to mitigate the COVID-19 pandemic. Some urban communities may be more vulnerable to rat infestations based on their location, housing, and income. For residents in small apartment buildings in restaurant-dense areas, proactive support may be needed in the form of rat abatement or information about rats. Targeted support may help minimize rat-associated risks such as zoonotic pathogen exposure or restricted use of outdoor spaces while staying at home. Such measures will help protect public health in cities struggling with rat infestations and the COVID-19 pandemic around the world.

## Data Availability

A summarized version of the datasets used and/or analyzed during the current study are available from the corresponding author on reasonable request.

## Declarations

### Ethics approval and consent to participate

This research involving human subjects was performed in accordance with the Declaration of Helsinki. The Lincoln Park Zoo Institutional Review Board (reference number IRB00009278) approved our informed consent protocol and deemed this study exempt from the requirements of 45 CFR 46 as per 45 CFR 46.104(d)(2) of U.S. Department of Health and Human Services regulations.

### Availability of data and materials

A summarized version of the datasets used and/or analysed during the current study are available from the corresponding author on reasonable request.

### Author’ contributions

MHM, KB, SBM, and DG designed the study. MHM led the quantitative analysis with assistance from all authors. MHM, KB, JB, DM, and PW contributed to the thematic analysis. MHM wrote the manuscript with assistance from all authors.

## Acknowledgments

We wish to thank the survey participants for sharing their experiences and the city officials and community organizations who shared our survey. We also thank Lydia Hopper, Lisa Hyatt, Elizabeth Lehrer, Cria Kay, Mason Fidino, and Kim Fake for their help with survey refinement and the volunteers who piloted our survey. Icons used in Figure 4 are from the Noun Project and created by Adrien Coquet, iconosphere, Maxim Kulikov, Alice Design, Vectors Point, Daryl Vandemont, Guilherme Furtado, and Pavel Pavlov.

## Supplementary Files

Supplementary File 1: Survey questionnaire developed for this study.

Supplementary File 2: Supplemental tables containing model outputs (Tables S1, S2, S3, S4) and a figure showing the relationship between the change in rat sightings and sightings of rat feces in the home (Figure S1).

