## Supplementary file 1 for "“I don’t feel safe sitting in my own yard”: Chicago resident experiences with urban rats during a COVID-19 stay-at-home order"

Corresponding author:

Name: Maureen H. Murray

### Urban Wildlife Institute

THE CHICAGO RAT PROJECT

#### Chicago Rat Project- Stay-at-Home Survey

##### Informed consent

**The Urban Wildlife Institute at Lincoln Park Zoo is continuing to learn as much as we can about Chicago's urban wildlife landscape as our communities navigate COVID-19 together. If you have time and are able, we would appreciate any insights you are able to provide via this survey. And, we understand if you aren't able to participate. Hopefully, we'll learn a little bit about our city and its non-human residents in this challenging environment.**

**Please only take this survey once, even if you have received it from multiple sources.**

**The Chicago Rat Project is studying how rats affect people in neighborhoods all across Chicago. We are asking for your help by participating in a brief survey that will help us learn from your experiences with rats. Your answers will help us understand rat issues in Chicago and how rats affect public perception of urban wildlife.**

**This survey is anonymous and it will take about 20 minutes. We will use your answers to write a scientific report. This project has been reviewed and approved by the Lincoln Park Zoo Institutional Review Board.**

**Benefits You will not benefit directly from this study. We hope our results will help reduce rats in Chicago to improve public health.**

**Risks Some of the survey questions may be uncomfortable to answer. For example, we will have images of rats and rat droppings. We will ask about rat droppings in your home and we will also ask if you are afraid when you see a rat. You can choose not to answer any question, for any reason.**

**Participation *Participating in this survey is completely voluntary.* You are free not to participate, to end the survey at any time for any reason, or to refuse to answer any question.**

**Confidentiality We will record your age and gender, but no other information about you. If you choose to provide your contact at the end of the survey, it will not be stored with your responses and will not be shared with any other party.**

**Persons to Contact If you have questions about this study, please contact Dr. Maureen Murray at. If you would like to talk with someone other than the researchers to discuss problems or concerns or to discuss your rights as a research participant, you may contact the Lincoln Park Zoo Institutional Review Board at.**

**Consent If you are willing to participate in this study, please check the boxes below. By filling out this survey, you are providing consent for us to use this data in our study. Please only fill out this survey if you are over 18 years of age and have lived in your residence for more than six months. Please note that checked boxes indicate that:**

**1. You consent to participate in this study**

**2. You understand that participation in this study is entirely voluntary and that you may refuse to participate or withdraw from the study at any time without penalty**

**3. You have received a copy of this consent form for your own records.**

* 1. I confirm that I am over 18 years old

- ◯ Yes
- ◯ No

* 2. I have lived in this property for more than **6 months**

- ◯ Yes
- ◯ No

Thank you for your help. Please keep this information in case you have questions later and want to contact us.

LINCOLN PARK ZOO

### Urban Wildlife Institute

THE CHICAGO RAT PROJECT

#### Chicago Rat Project- Stay-at-Home Survey

##### How to identify rats and their signs

Norway or brown rats are bigger and longer in shape than mice.

| | | |
|---|---|---|
| House mouse | 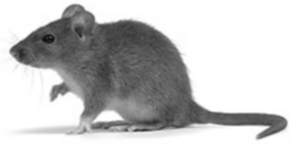 | 0.5 – 1 oz<br>3 – 4” long (body)<br>5.5 – 7.5” long<br>(with tail) |
| Dollar bill | 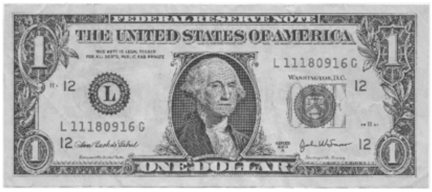 | 6.14” long |
| Brown/Norway rat | 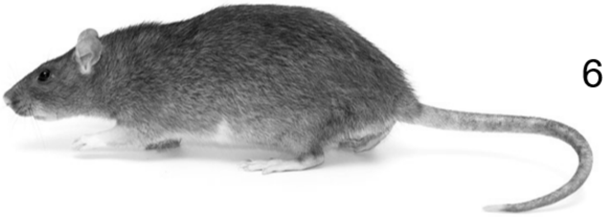 | 7 – 18 oz<br>6 – 11” long (body)<br>13 – 18” long<br>(with tail) |

Rat droppings are larger than grains of rice, which are larger than mouse droppings.

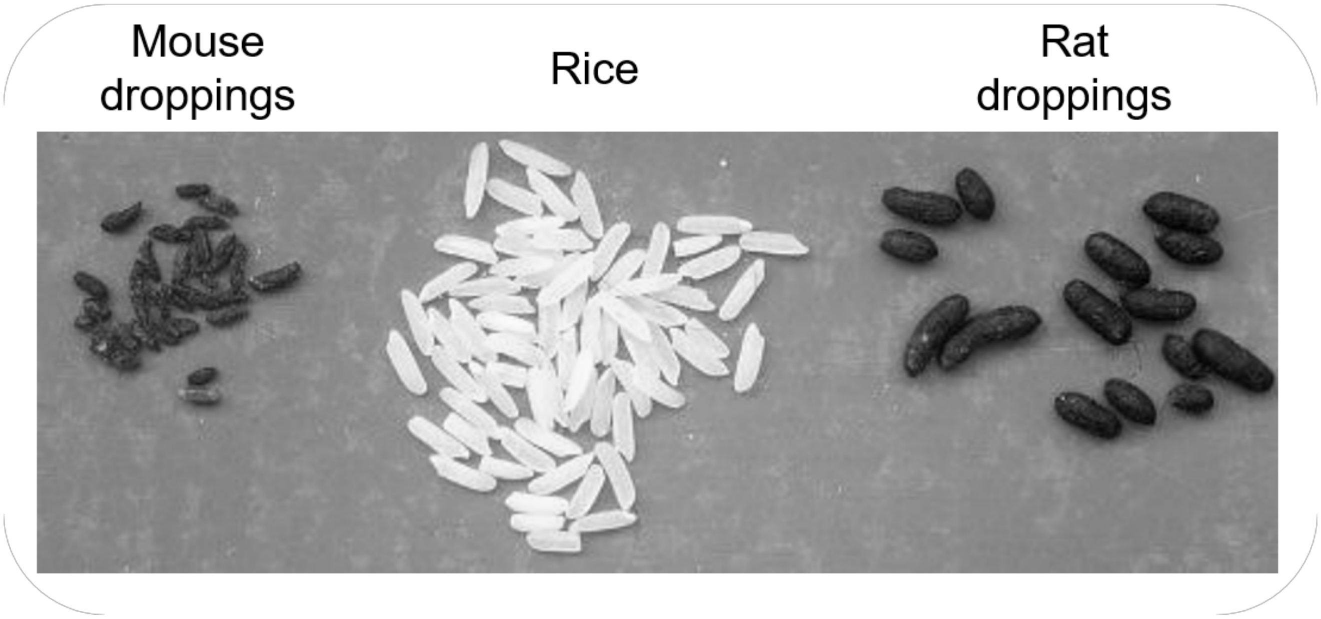


Rats dig holes or burrows in the ground, like this one next to a dumpster.

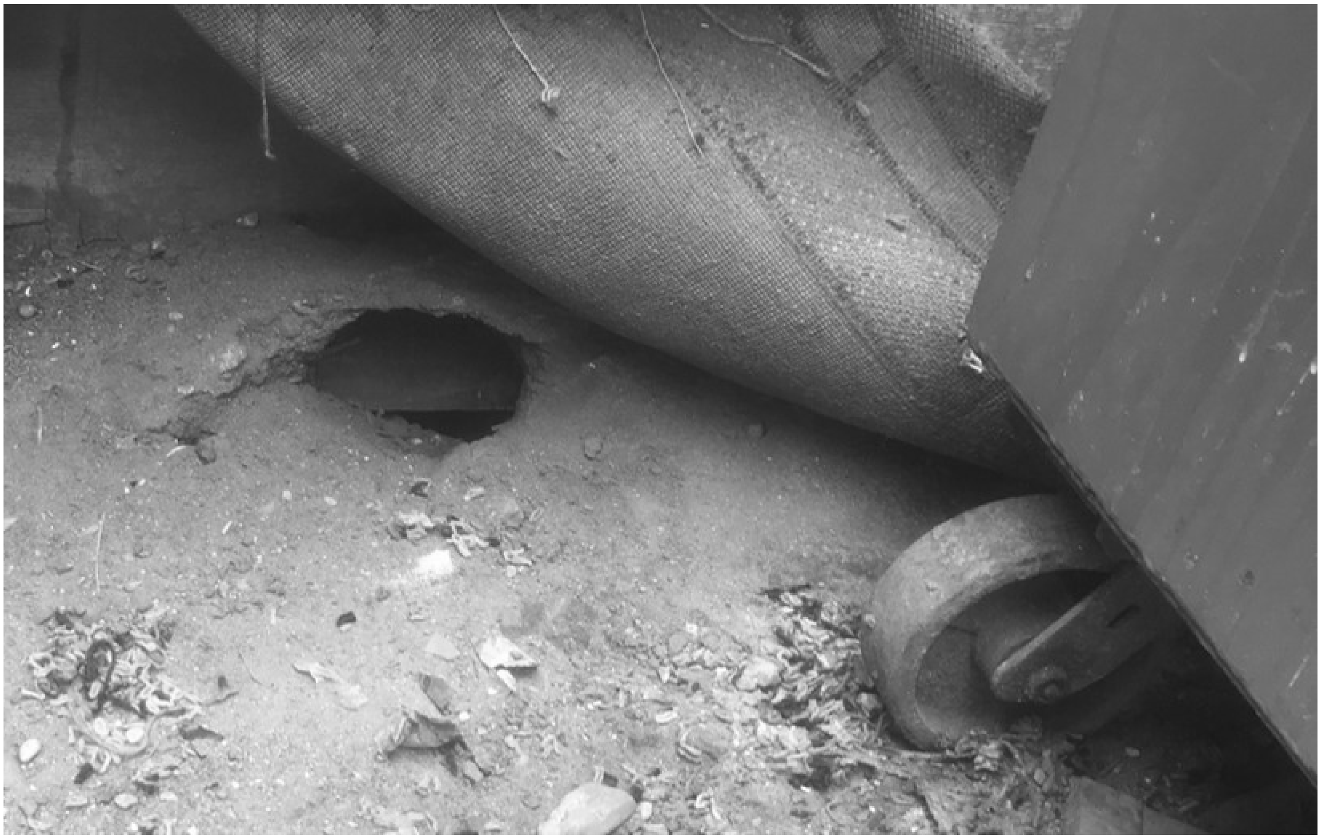

LINCOLN PARK ZOO

### Urban Wildlife Institute

THE CHICAGO RAT PROJECT

#### Chicago Rat Project- Stay-at-Home Survey

##### Experiences with rats

***These questions tell us where, when, and how often Chicago residents see rats.***

3. Thinking over the **past month**, how often have you seen a rat in each of these places?

| | Never | Rarely (Less than weekly) | Frequently (Weekly) | Daily or almost daily |
|---|---|---|---|---|
| In your neighborhood | ○ | ○ | ○ | ○ |
| On your block | ○ | ○ | ○ | ○ |
| In or around your home/building | ○ | ○ | ○ | ○ |

4. Compared to **a month ago**, how often are you seeing rats now?

| | Fewer rats | About the same | More rats | I do not see any rats here |
|---|---|---|---|---|
| In your neighborhood | ○ | ○ | ○ | ○ |
| On your block | ○ | ○ | ○ | ○ |
| In or around your home | ○ | ○ | ○ | ○ |

5. Compared to previous years, how often are you seeing rats this year?

○ Fewer rats

○ About the same

○ More rats

6. When you think about rats, what words come to mind?

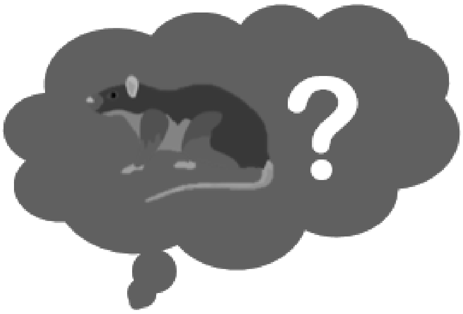

#### Chicago Rat Project- Stay-at-Home Survey

##### Attitudes about rats

***This question tells us what Chicago residents think about rats.***

7. For each of the following statements, please tell us how much you agree or disagree:

| | Strongly disagree | Disagree | Neither agree nor disagree | Agree | Strongly agree |
|---|---|---|---|---|---|
| Seeing rats bothers me | ◯ | ◯ | ◯ | ◯ | ◯ |
| Rats are smart | ◯ | ◯ | ◯ | ◯ | ◯ |
| Rats cause property damage | ◯ | ◯ | ◯ | ◯ | ◯ |
| Rats are cute | ◯ | ◯ | ◯ | ◯ | ◯ |
| Rats spread disease | ◯ | ◯ | ◯ | ◯ | ◯ |
| Rats are dangerous | ◯ | ◯ | ◯ | ◯ | ◯ |
| Rats are scary | ◯ | ◯ | ◯ | ◯ | ◯ |
| Rats are unclean | ◯ | ◯ | ◯ | ◯ | ◯ |
| I think about rats even when I don’t see them | ◯ | ◯ | ◯ | ◯ | ◯ |
| I am concerned about rat infestations | ◯ | ◯ | ◯ | ◯ | ◯ |
| I am more concerned about rats than I was a month ago | ◯ | ◯ | ◯ | ◯ | ◯ |

8. Please tell us how concerned you are about these potential consequences of rats:

| | Not at all | A little | Somewhat | Very |
|---|---|---|---|---|
| Property Damage | ◯ | ◯ | ◯ | ◯ |
| Disease | ◯ | ◯ | ◯ | ◯ |
| Safety | ◯ | ◯ | ◯ | ◯ |
| Environmental Damage | ◯ | ◯ | ◯ | ◯ |

9. Have you ever owned a pet rat?

◯ Yes ◯ No

#### Chicago Rat Project- Stay-at-Home Survey

##### Rat Control

***These questions tell us about how Chicago residents control rats and what they think about rat management.***

10. Over the **past month**, where have you found information about rats? Please check all that apply.

- ☐ Television
- ☐ Friends or family
- ☐ City of Chicago posters
- ☐ City of Chicago website
- ☐ Other websites
- ☐ Neighbors
- ☐ Other (please specify)
- ☐ Other community members
- ☐ Pest professionals
- ☐ Social media
- ☐ Radio
- ☐ I have not looked for information about rats

11. Thinking over the **past month**, how often have you done any of these activities?

| | Never | Rarely (Less than weekly) | Frequently (Weekly) | Daily or almost daily |
|---|---|---|---|---|
| Called 311 to report a rat | ○ | ○ | ○ | ○ |
| Set snap traps to kill rats | ○ | ○ | ○ | ○ |
| Used rat poison | ○ | ○ | ○ | ○ |
| Replaced garbage cans because of rat holes | ○ | ○ | ○ | ○ |
| Covered rat holes | ○ | ○ | ○ | ○ |
| Called a pest professional about rats | ○ | ○ | ○ | ○ |
| Cleaned up building or yard to prevent rats | ○ | ○ | ○ | ○ |

12. Compared to **a month ago**, how often have you needed to use rodent control?

- Did not use rodent control
- Less often
- About the same
- More often

13. Compared to **previous years**, how often are you using rodent control this year?

- I used less compared to last year
- About the same as last year
- I used more than last year

14. Thinking over the **past month**, for each of the following statements, please tell us how much you agree or disagree:

| | Strongly disagree | Disagree | Neither agree nor disagree | Agree | Strongly agree |
|---|---|---|---|---|---|
| The City of Chicago is doing a good job at getting rid of rats | | | | | |
| The rat complaints in my neighborhood are taken seriously | | | | | |
| I have enough information and resources to control rats on my property | | | | | |
| My neighbors do a good job getting rid of rats | | | | | |
| I do everything I can to get rid of rats | | | | | |

15. Please rank the following people in order of who should be responsible for rat control, with 1 being the most responsible and 3 the least responsible

Landlords/Building owners

Renters

The government

16. In your opinion, what do you think is the best way to manage rats in your neighborhood? Please rank the actions below in order with 1 being the most effective, to 5 being the least effective.

Cleaning up the garbage

Putting out rat poison bait

Putting out rat traps

Filling in burrows

Repair homes

17. Are there other ways you think would help manage rats? If yes, please explain.

LINCOLN PARK ZOO

Urban Wildlife Institute

THE CHICAGO RAT PROJECT

#### Chicago Rat Project- Stay-at-Home Survey

##### Your neighborhood

***These questions help us understand which neighborhoods or housing types need more help dealing with rats.***

18. Do you rent or own your residence?

- ◯ Rent
- ◯ Own

19. What type of housing do you live in?

- ◯ Single-family home
- ◯ Townhouse
- ◯ Apartment building with 2 – 4 units
- ◯ Apartment building with 5 – 9 units
- ◯ Apartment building with 10 – 19 units
- ◯ Apartment building with 20 – 49 units
- ◯ Apartment building with over 50 units

20. What neighborhood do you live in?

[ ]

21. Optional: What is the closest major street intersection to your block? (example: N Clark St and W Armitage Ave)

[ ]

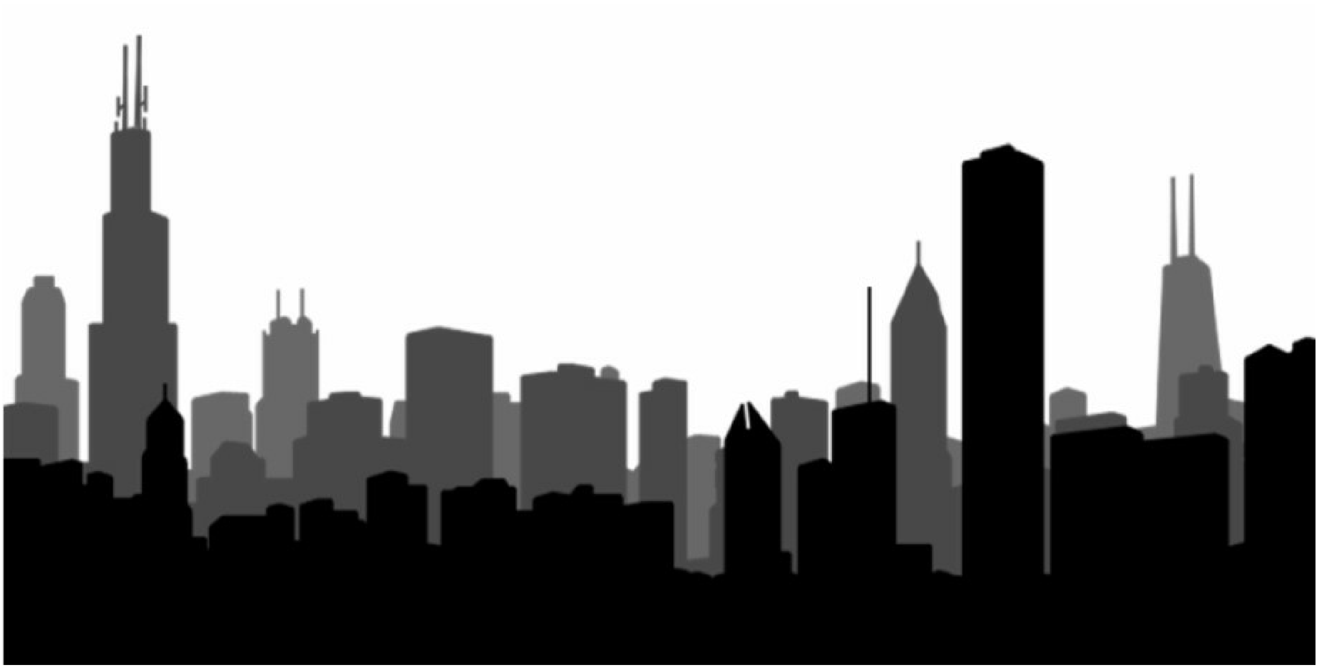

22. Thinking over the **past month**, for each of the following, please tell us if this is “not a problem”, “somewhat of a problem”, or “a big problem” on your block:

| | Not a problem | Somewhat of a problem | A big problem |
|---|---|---|---|
| Rats | ◯ | ◯ | ◯ |
| Vacant housing | ◯ | ◯ | ◯ |
| Vandalism | ◯ | ◯ | ◯ |
| Trash in the streets | ◯ | ◯ | ◯ |
| Groups of teenagers hanging out in the street | ◯ | ◯ | ◯ |
| People selling drugs | ◯ | ◯ | ◯ |
| People getting robbed or beat up | ◯ | ◯ | ◯ |

### Urban Wildlife Institute

LINCOLN PARK ZOO

THE CHICAGO RAT PROJECT

#### Chicago Rat Project- Stay-at-Home Survey

##### Rats and your health

***These questions help us understand how rats might be related to health for Chicago residents.***

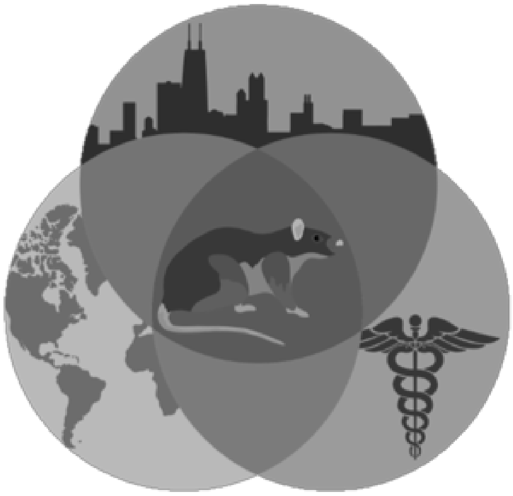

23. If you currently have rat problems or had them in the past, how does that make you feel? Please describe your thoughts and feelings in your own words.

24. Over the **past month**, how frequently have you seen these signs of rats in your home/building?

| | Never | Rarely (Less than weekly) | Frequently (Weekly) | Daily or almost daily |
|---|---|---|---|---|
| Rat-chewed walls, wires, or pipes | ○ | ○ | ○ | ○ |
| Rat-chewed food containers | ○ | ○ | ○ | ○ |
| Noticed a strong smell of rats or rat urine | ○ | ○ | ○ | ○ |
| Rat droppings/feces | ○ | ○ | ○ | ○ |

25. Do you take off your shoes when you come indoors?

◯ Most of the time ◯ Sometimes ◯ Rarely

26. Over the **past month**, how often have you experienced any of these events?

| | Never | Rarely (Less than weekly) | Frequently (Weekly) | Daily or almost daily |
|---|---|---|---|---|
| Touched a rat (alive or dead) | ◯ | ◯ | ◯ | ◯ |
| Touched rat droppings | ◯ | ◯ | ◯ | ◯ |
| Bitten by a rat | ◯ | ◯ | ◯ | ◯ |

27. Compared to a **month ago**, how often do you have the following rat issues?

| | Less often | About the same | More Often | Never |
|---|---|---|---|---|
| Saw chewed objects | ◯ | ◯ | ◯ | ◯ |
| Saw rat droppings | ◯ | ◯ | ◯ | ◯ |
| Touched rat droppings | ◯ | ◯ | ◯ | ◯ |
| Touched a rat | ◯ | ◯ | ◯ | ◯ |
| Bitten by a rat | ◯ | ◯ | ◯ | ◯ |

28. If you experienced any of the events in the previous question, did you have any health issues from coming into contact with a rat or rat droppings?

◯ Yes ◯ No

If yes, please explain:

29. Did you see a doctor or go to a health clinic because of this rat related health issue?

◯ Yes

◯ No

If yes, please explain

LINCOLN PARK ZOO

### Urban Wildlife Institute

THE CHICAGO RAT PROJECT

#### Chicago Rat Project- Stay-at-Home Survey

##### Urban wildlife

***These questions help us understand what Chicago residents think and experience about other urban wildlife (wild animals that live in Chicago).***

30. What words come to mind when you think about wildlife in cities?

31. For each of the following statements about wildlife management (removing animals to control the population), please tell us how much you agree or disagree:

| | Strongly disagree | Disagree | Neither agree nor disagree | Agree | Strongly agree |
|---|---|---|---|---|---|
| Humans should manage wild animal populations so that humans benefit | ○ | ○ | ○ | ○ | ○ |
| If a type of animal is common, we should manage those animals to improve human lives | ○ | ○ | ○ | ○ | ○ |
| It is acceptable to kill animals for management, if that type of animal is common | ○ | ○ | ○ | ○ | ○ |

32. For each of the following statements about wildlife rights, please tell us how much you agree or disagree

| | Strongly Disagree | Disagree | Neither Agree or Disagree | Agree | Strongly Agree |
|---|---|---|---|---|---|
| Killing wildlife is wrong because animals have a right to exist | ○ | ○ | ○ | ○ | ○ |
| Animals should have rights similar to the rights of humans | ○ | ○ | ○ | ○ | ○ |
| The rights of wildlife are more important than the management of wildlife | ○ | ○ | ○ | ○ | ○ |

LINCOLN PARK ZOO
### Urban Wildlife Institute
THE CHICAGO RAT PROJECT

#### Chicago Rat Project- Stay-at-Home Survey

##### Urban wildlife

***These questions tell us how often residents in different neighborhoods see urban wildlife.***

33. These are some common mammals in Chicago. **In the past month**, which of the following mammals have you seen in your neighborhood? Please click all images that apply

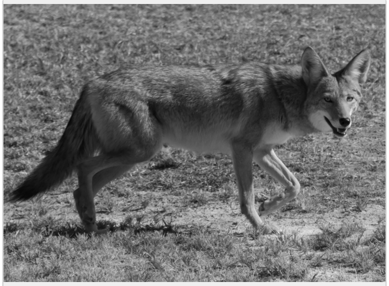
Coyote

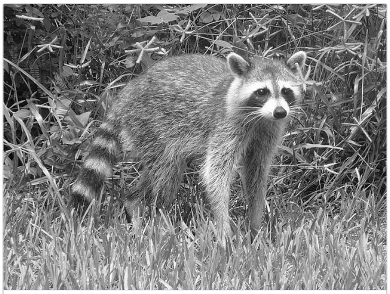
Raccoon

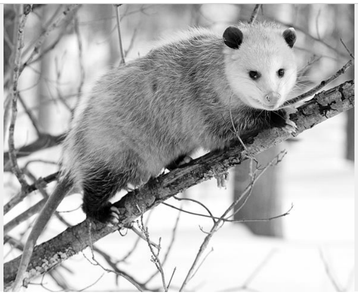
Opossum

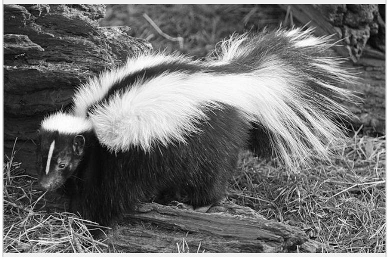
Skunk

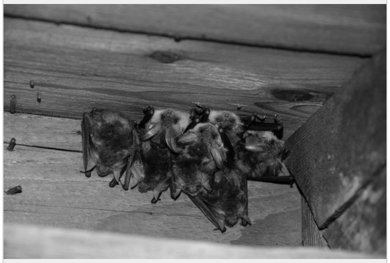
Bat (any kind)

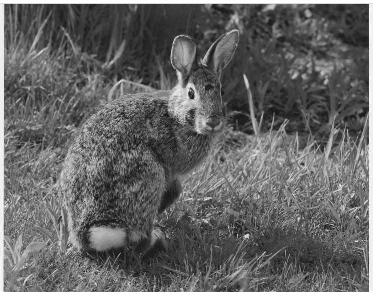
Rabbit

34. These are some common birds in Chicago. **In the past month**, which of the following birds have you seen in your neighborhood? Please click all images that apply.

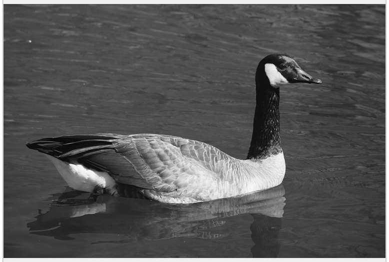

Canada goose

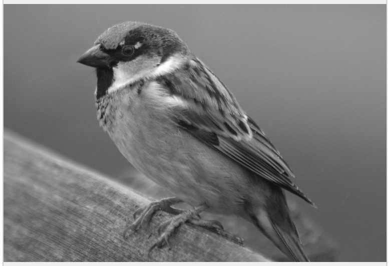

House sparrow

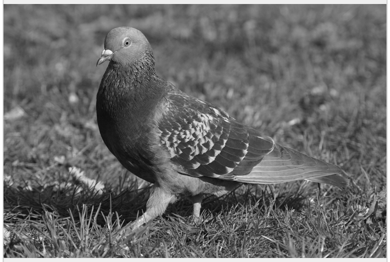

Pigeon

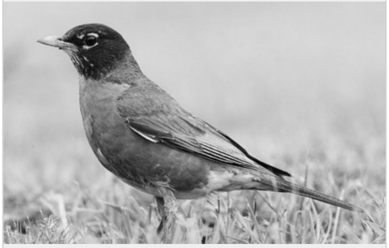

Robin

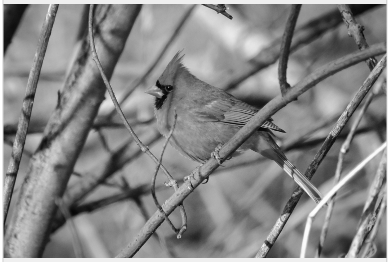

Cardinal

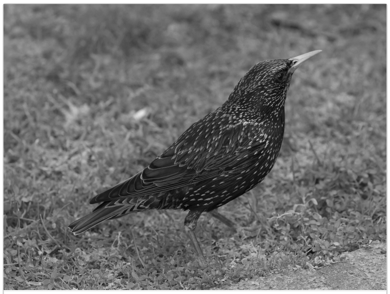

Starling

### Chicago Rat Project- Stay-at-Home Survey

#### Attitudes about urban wildlife

***These questions tell us what Chicago residents think about urban wildlife (wild animals in the city).***

35. For each of the following statements about urban parks, please tell us how much you agree or disagree:

| | Strongly disagree | Disagree | Neither agree nor disagree | Agree | Strongly agree |
|---|---|---|---|---|---|
| I enjoy watching wildlife when I visit urban parks | ◯ | ◯ | ◯ | ◯ | ◯ |
| One of the reasons I go to urban parks is to see wildlife | ◯ | ◯ | ◯ | ◯ | ◯ |
| I am interested in learning about the wildlife that live in Chicago's parks | ◯ | ◯ | ◯ | ◯ | ◯ |

36. For each of the following statements about urban wildlife around your home or community, please tell us how much you agree or disagree:

| | Strongly disagree | Disagree | Neither agree nor disagree | Agree | Strongly agree |
|---|---|---|---|---|---|
| I enjoy seeing wildlife around my home | ◯ | ◯ | ◯ | ◯ | ◯ |
| I'm interested in making the area around my home attractive to wildlife | ◯ | ◯ | ◯ | ◯ | ◯ |
| I enjoy seeing wildlife in my neighborhood | ◯ | ◯ | ◯ | ◯ | ◯ |

37. For each of the following statements, please tell us how much you agree or disagree:

| | Strongly Disagree | Disagree | Neither Agree or Disagree | Agree | Strongly Agree |
|---|---|---|---|---|---|
| In the past month, I have noticed more wildlife around my home | ◯ | ◯ | ◯ | ◯ | ◯ |
| In the past month, I have noticed more wildlife in my neighborhood | ◯ | ◯ | ◯ | ◯ | ◯ |
| In the past month, I have thought about wildlife in cities more often than usual | ◯ | ◯ | ◯ | ◯ | ◯ |

LINCOLN PARK ZOO®

### Urban Wildlife Institute

THE CHICAGO RAT PROJECT

#### Chicago Rat Project- Stay-at-Home Survey

##### Please tell us about yourself

***This information will remain confidential and anonymous.***

38. What is your age group?

- ◯ 18-24
- ◯ 25-34
- ◯ 35-44
- ◯ 45-54
- ◯ 55-64
- ◯ 65+

39. What is your gender?

- ◯ Female
- ◯ Male
- ◯ Non-binary
- ◯ Prefer not to say

40. Do you have any children under 18 living at home?

- ◯ Yes
- ◯ No

41. How many people live in your household? Please leave blank if you prefer not to say.

____________________

42. Thinking over the **past month**, how much time do you spend outdoors in your neighborhood in a typical week?

- ◯ Under 2 hours
- ◯ Between 2-6 hours
- ◯ Between 7 - 14 hours
- ◯ More than 14 hours

43. How did you hear about this survey? (Please select all that apply)

- ☐ Alderman Newsletter
- ☐ E-mail from community organization
- ☐ Shared by a friend
- ☐ Social Media
- ☐ I am a Lincoln Park Zoo volunteer
- ☐ I am a Field Museum volunteer
- ☐ Other (please specify)

#### Thank you for filling out our survey!

**This is the first phase of a multi-year study about rats in Chicago.
If you are interested in project updates,
taking a follow-up survey, or to
get involved in community-based research on rat management,
please leave your contact information below.**

**Your contact information will remain confidential and will be kept separate from your survey answers.**

**We would appreciate your feedback, which you can leave on the next page. Thank you!**

44. Contact information

**Name**

**Email Address**

**Phone Number**

LINCOLN PARK ZOO

### Urban Wildlife Institute

THE CHICAGO RAT PROJECT

#### Chicago Rat Project- Stay-at-Home Survey

##### Pilot Survey Feedback

***Please let us know what you thought of this survey. We appreciate your feedback!***

45. Were there any questions that you found confusing?

46. Were there any questions you were not comfortable answering?

47. What did you think of the survey length?

- ◯ Too short
- ◯ About the right length
- ◯ Too long

48. Were there any questions you wished we had asked?
