## Supplementary file 2 for "“I don’t feel safe sitting in my own yard”: Chicago resident experiences with urban rats during a COVID-19 stay-at-home order"

Corresponding author:

Name: Maureen H. Murray

Table S1: Demographic characteristics of survey respondents in Chicago, Illinois, USA during the spring 2020 stay-at-home order.

| Characteristic | Survey respondents  (n = 835) | Chicago 2010 census |
| --- | --- | --- |
| **Age class** |  |  |
| 18-24 | 3.47% | 11.20% |
| 25-34 | 14.61% | 19.10% |
| 35-44 | 18.80% | 14.00% |
| 45-54 | 16.88% | 12.60% |
| 55-64 | 14.13% | 9.80% |
| 65+ | 12.69% | 10.30% |
| NA | 19.40% | - |
| **Gender** |  |  |
| Female | 54.25% | 51.57% |
| Male | 24.55% | 48.53% |
| Non-Binary | 0.60% | NA |
| Prefer not to say | 1.08% | NA |
| NA | 19.52% | - |
| **Property Ownership** | |  |
| Own | 66.22% | 44.91% |
| Rent | 22.99% | 55.01% |
| NA | 10.79% |  |

Table S2: Ordinal regression output for variables associated with a change in rat sightings in or around the respondent’s home during the quarantine period.

| Variable | 𝛃 | Std. Error | t value | p value |
| --- | --- | --- | --- | --- |
| Restaurants | 0.08 | 0.09 | 0.83 | 0.40 |
| Gender (Male) | -0.01 | 0.18 | -0.04 | 0.97 |
| Age (Linear) | 0.16 | 0.37 | 0.43 | 0.67 |
| Age (Quadratic) | -0.45 | 0.31 | -1.44 | 0.15 |
| Age (Cubic) | -0.42 | 0.25 | -1.70 | 0.09 |
| Age (^4) | -0.02 | 0.20 | -0.11 | 0.91 |
| Age (^5) | -0.16 | 0.18 | -0.87 | 0.38 |
| Time outside (Linear) | 0.49 | 0.23 | 2.18 | 0.03 |
| Time outside (Quadratic) | 0.11 | 0.19 | 0.59 | 0.56 |
| Time outside (Cubic) | -0.01 | 0.15 | -0.04 | 0.97 |
| Renter | 0.01 | 0.25 | 0.05 | 0.96 |
| Housing (large multi-unit) | -0.77 | 0.26 | -2.96 | 3.08 x 10-3 |
| Housing (small multi-unit) | 0.50 | 0.20 | 2.55 | 0.01 |
| Median household income | 0.08 | 0.09 | 0.87 | 0.38 |
| 311 complaints | 0.08 | 0.10 | 0.73 | 0.47 |

Table S3: Logistic regression output for variables hypothesized to be associated with the likelihood of a survey respondent calling 311 during quarantine.

| Variable | 𝛃 | Std. Error | z value | p value |
| --- | --- | --- | --- | --- |
| Intercept | -1.36 | 0.33 | -4.11 | 3.90 x 10-5 |
| Change in rat sightings at home (Linear) | 1.67 | 0.45 | 3.72 | 2.03 x 10-4 |
| Change in rat sightings at home (Quadratic) | -0.80 | 0.39 | -2.03 | 0.04 |
| Change in rat sightings at home (Cubic) | 0.31 | 0.32 | 0.98 | 0.33 |
| Change in concern about rats (Linear) | 2.01 | 0.42 | 4.75 | 2.00 x 10-6 |
| Change in concern about rats (Quadratic) | 0.04 | 0.37 | 0.12 | 0.91 |
| Change in concern about rats (Cubic) | -0.06 | 0.32 | -0.18 | 0.86 |
| Change in concern about rats (^4) | -0.16 | 0.27 | -0.59 | 0.55 |
| Information about rats (Linear) | 0.66 | 0.41 | 1.60 | 0.11 |
| Information about rats (Quadratic) | 1.27 | 0.37 | 3.45 | 5.63 x 10-4 |
| Information about rats (Cubic) | 0.21 | 0.29 | 0.73 | 0.47 |
| Information about rats (^4) | 0.34 | 0.24 | 1.41 | 0.16 |
| Gender (Male) | -0.30 | 0.26 | -1.14 | 0.26 |
| Children (Yes) | 0.30 | 0.30 | 1.02 | 0.31 |
| Age (Linear) | 0.28 | 0.73 | 0.39 | 0.70 |
| Age (Quadratic) | -0.11 | 0.65 | -0.17 | 0.86 |
| Age (Cubic) | 0.03 | 0.48 | 0.07 | 0.95 |
| Age (^4) | -0.12 | 0.35 | -0.34 | 0.73 |
| Age (^5) | 0.11 | 0.27 | 0.41 | 0.68 |
| Renter | -1.13 | 0.40 | -2.78 | 0.01 |
| Median household income | -0.11 | 0.12 | -0.96 | 0.34 |

Table S4: Ordinal regression output for variables hypothesized to be associated with the likelihood of a survey respondent calling a pest control professional during quarantine.

| Variable | 𝛃 | Std. Error | z value | p value |
| --- | --- | --- | --- | --- |
| Intercept | -1.08 | 0.30 | -3.58 | 3.42 x 10-4 |
| Change in rat sightings at home (Linear) | 0.81 | 0.36 | 2.26 | 0.02 |
| Change in rat sightings at home (Quadratic) | -0.68 | 0.32 | -2.13 | 0.03 |
| Change in rat sightings at home (Cubic) | 0.35 | 0.28 | 1.25 | 0.21 |
| Change in concern about rats (Linear) | 1.34 | 0.37 | 3.57 | 3.52 x 10-4 |
| Change in concern about rats (Quadratic) | 0.06 | 0.33 | 0.20 | 0.84 |
| Change in concern about rats (Cubic) | -0.34 | 0.32 | -1.07 | 0.29 |
| Change in concern about rats (^4) | 0.21 | 0.27 | 0.77 | 0.44 |
| Information about rats (Linear) | 0.23 | 0.40 | 0.57 | 0.57 |
| Information about rats (Quadratic) | 1.17 | 0.36 | 3.27 | 1.07 x 10-3 |
| Information about rats (Cubic) | 0.09 | 0.28 | 0.33 | 0.74 |
| Information about rats (^4) | -0.01 | 0.24 | -0.03 | 0.98 |
| Gender (Male) | -0.08 | 0.25 | -0.30 | 0.77 |
| Children (Yes) | 0.08 | 0.29 | 0.29 | 0.77 |
| Age (Linear) | -0.12 | 0.74 | -0.16 | 0.87 |
| Age (Quadratic) | 0.17 | 0.65 | 0.26 | 0.80 |
| Age (Cubic) | -0.09 | 0.48 | -0.18 | 0.86 |
| Age (^4) | 0.04 | 0.36 | 0.11 | 0.91 |
| Age (^5) | 0.27 | 0.27 | 1.01 | 0.31 |
| Renter | -1.72 | 0.47 | -3.64 | 2.73 x 10-4 |
| Median household income | 0.07 | 0.11 | 0.62 | 0.54 |

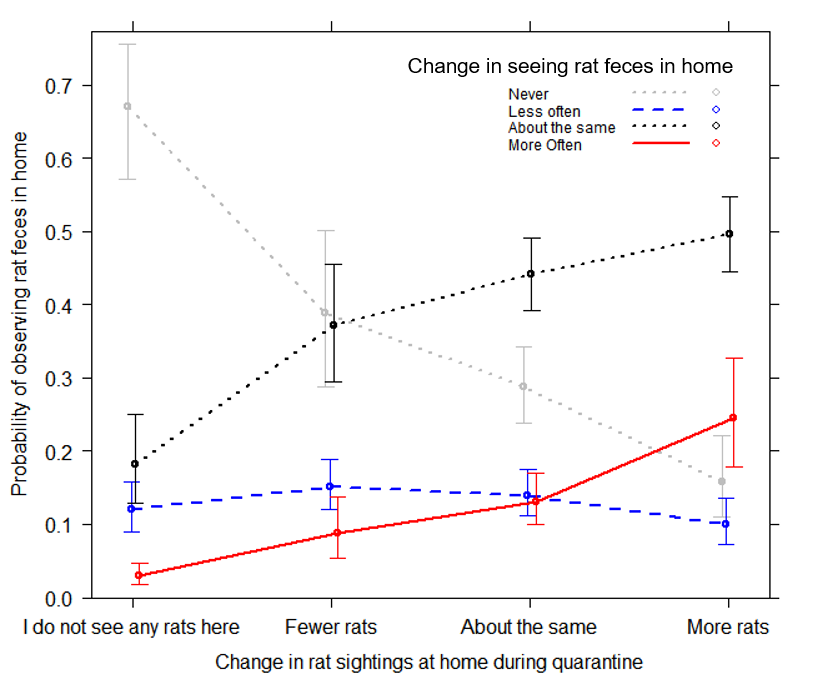

Figure S1: Relationship between survey respondents observing rats and rat feces in their home during quarantine.
